# The COVID-19 Suffolk Events Toolkit (C-SET): A structured approach to conducting COVID-secure events

**DOI:** 10.1101/2020.10.23.20218412

**Authors:** Avirup Chowdhury, Terezija Zermanos, Xinming Yu, Padmanabhan Badrinath

## Abstract

**Objectives:** To develop a toolkit to provide a structured framework for assessing whether large events are COVID-secure.

**Study design:** Cross-sectional reporting on the development of the COVID-19 Suffolk Events Toolkit (C-SET).

**Methods:** The toolkit was developed through an iterative process of writing, discussion, and modification, drawing upon national UK guidance. Pilot cases and a contemporaneous consultation with experts were also undertaken, and C-SET was revised in line with the findings.

**Results:** C-SET offers a robust, structured approach to identifying areas of concern and potential mitigations, and collates and synthesises current national UK guidance. A flexible, modular design allows it to provide high-level input across all types of event, and allow modification to suit the needs of individual organisations. It is applicable to a variety of users, including event organisers in need of a structure, or bodies whose role is to appraise such events. It provides a semi-quantitative summary output, and in conjunction with the detailed links to guidance in the framework, allows development of clear, actionable recommendations to tangibly improve the quality of events.

**Conclusions:** A necessary trade-off exists between COVID-related health risks and wider risks due to economic downturn, and facilitating large events in a way that minimises COVID transmission risk may offer a way to balance these competing priorities. C-SET was developed to provide a structured approach to identifying areas of improvement across key risk domains, and suggests potential mitigations that are underpinned by national UK guidance. Use of this toolkit may aid event organisers in developing COVID-secure events, and aid regulatory bodies in suggesting actionable recommendations through generation of a summary report of areas of concern.

## Introduction

Since the beginning of the COVID-19 global pandemic, the negative impact of COVID-19 transmission, primarily in the form of increased morbidity and mortality, has been well-established^1^. However, there are also negative health outcomes in COVID-19 resulting from socioeconomic disadvantage^2,3^, itself more likely as a result of prolonged economic recession, as well as negative health effects of prolonged lockdown (such as in mental health outcomes^3–5^ and the excess deaths due to resource allocation towards COVID-19^1,6^). This has given rise to a tension between the economic and health advantages of opening the economy and the negative health consequences of a potential increase in COVID-19 transmission. This is most evident when considering mass gatherings, which may have a positive impact upon the economy, but a potential increase in COVID-19 transmission risk.

A mass gathering is defined by WHO as “a planned or spontaneous event that gathers substantial numbers of attendees who might strain the health planning and response capacities of the host community, city, or country”^7^. These are key dilemmas for health authorities and governments^8^: while mass gatherings are important for economic reasons, they often bring risks when potentially thousands of people living in areas with differing COVID-19 prevalence travel long distances to attend a single event. In addition, mass gatherings have historically been the source of infectious disease spread globally^7^; this continues to be true for COVID-19 in the UK^9^ and beyond^10^. Furthermore, there is potential for outbreaks when people come together as shown by the recent example of students travelling to universities for the start of the new academic year^11,12^.

The evidence base for infectious disease transmission at mass gatherings is still developing^7,8,13^; this paper focuses specifically upon a subset of these, namely organised events (specifically to exclude informal or otherwise uncoordinated gatherings). Creation of so-called “COVID-secure” events (i.e. those with appropriate measures in place to minimise risks as far as feasible) may allow a positive trade-off between these opposing interests, and may allow leveraging of the positive effects of large events while minimising the negative COVID-19 health risks to the wider community^14^. It is the authors’ professional experience that there is a broad variation in size, scope and nature of the mitigations proposed by organisers of large gatherings; this has similarly been reported in literature^14^. This is not unexpected, given the variation in size, scope and resourcing of such events. Organisational documentation used to report these events to relevant bodies also tends to vary in quality and completeness.

The evidence base for COVID-19 is limited and evolving, including the relevant legislations. Both the organisers of events and the public bodies which work with them to ensure events are COVID-secure have limited resources, including time and expertise. A report from the Centre for Evidence-Based Medicine^15^ also concluded that mass gatherings are heterogeneous and require assessment on a case-by-case basis, potentially increasing the burden upon organisers and public bodies. Hence, there is an urgent need to develop a structured approach to this complex issue and supporting tools.

### COVID-19 Suffolk Events Toolkit

The COVID-19 Suffolk Events Toolkit (C-SET, appendices 1-3) has been created to facilitate a systematic appraisal of best practices to minimise risks arising as a consequence of large events during the COVID-19 pandemic, drawing upon national guidance to define key risk domains and incorporating work from other groups including the World Health Organisation^8^. The authors of this study, members of the Directorate of Public Health in Suffolk County Council, have developed this based on their experience in dealing with events in the COVID era. It has not been designed with a single user-group in mind, and may provide a guide for organisations planning COVID-secure events, or aid statutory bodies in appraising events and suggesting mitigations. It provides general principles which are applicable to all events, with additional sections for specific risk areas (e.g. accommodation, retail) which may not apply universally. It is the authors’ hope that this will be of interest to event organisers and public bodies.

## Methods

### Development of the Toolkit

The Toolkit was developed by the authors, who work in public health in a Local Authority. Framework structure was agreed through discussions between the authors. An iterative process of writing, discussion and modification was used to develop the framework, drawing upon national guidance.

It included any national guidance produced by the UK government and published on the GOV.UK website (https://www.gov.uk/) which may be relevant to large events (such as the “working safely during coronavirus” guidance). This guidance was chosen as it tended to be sufficiently detailed to produce general principles and mitigations, and can be considered the “gold standard” of policy. Third-party guidance tended to delve into more specific circumstances, and is suggested as an additional resource in the framework where relevant. Guidance which was relevant to individuals but not large groups (e.g. individual use of gyms/leisure facilities) was not included.

A list of guidelines from which this framework was developed is available the framework document (Appendix 1); however, this is not an exhaustive list of all guidance considered during the development process.

Part of the development process included undertaking pilot cases and a contemporaneous consultation with experts working in the front-line to ensure events are COVID-secure in Suffolk and beyond. These included senior colleagues from the Safety Advisory Groups (SAGs) of local authorities and Trading Standards. Two pilots were undertaken (one by one of the authors – XY – and the other by an external expert), testing the toolkit against documents provided from previous large events. All the consultees are also end-users of the toolkit. C-SET was revised in line with the comments from this consultation and findings from pilots. As part of the revision process based on the consultation and pilots, the checklist proforma was restructured through development of a modular design, and key risk domains in the framework were expanded and developed further.

### Scope of the Toolkit

This toolkit is suitable for use as an assessment framework for all large gatherings. It considers domains of risk which are common across all large gatherings (e.g. site access and crowd control, staff and workforce) and some which may be appropriate only to certain settings (e.g. accommodation). It is suitable for assessing the breadth of mitigations in place, and signposts users to the current national guidance in each instance.

Information on powers to impose conditions upon events or cancelling them altogether, and the legal frameworks underpinning this, are beyond the scope of C-SET. It also does not consider modifications required as part of event preparation (e.g. rehearsals for performing arts events, or construction of temporary stands ahead of outdoor sporting events).

## Results

### Components of the toolkit

C-SET consists of three documents:

1. Framework (Appendix 1): the evidence base for risk areas and mitigations.
2. Checklist (Appendix 2): a structured tool for examining all relevant dimensions of an event, providing a summary report.
3. User manual (Appendix 3): provides support for end-users completing the checklist.

### Structure of the framework

The framework summarises national guidance and groups it into ten key risk domains (Table 1); some of these are common to all large gatherings, whilst others might only be appropriate for certain events. Each domain has further sub-domains, and within these are individual considerations. A suggested mitigation, related question and links to guidance (with title and section number) are provided in each instance. For example, within the “ingress and egress” sub-domain (Appendix 1, Section 2.4), a key consideration is “one-way systems”. The suggested mitigation for this is to “use markings and introduce an accessible one-way flow at entry and exit points”, and is linked to the appropriate performing arts and visitor economy guidance for further details.

**Table 1.**
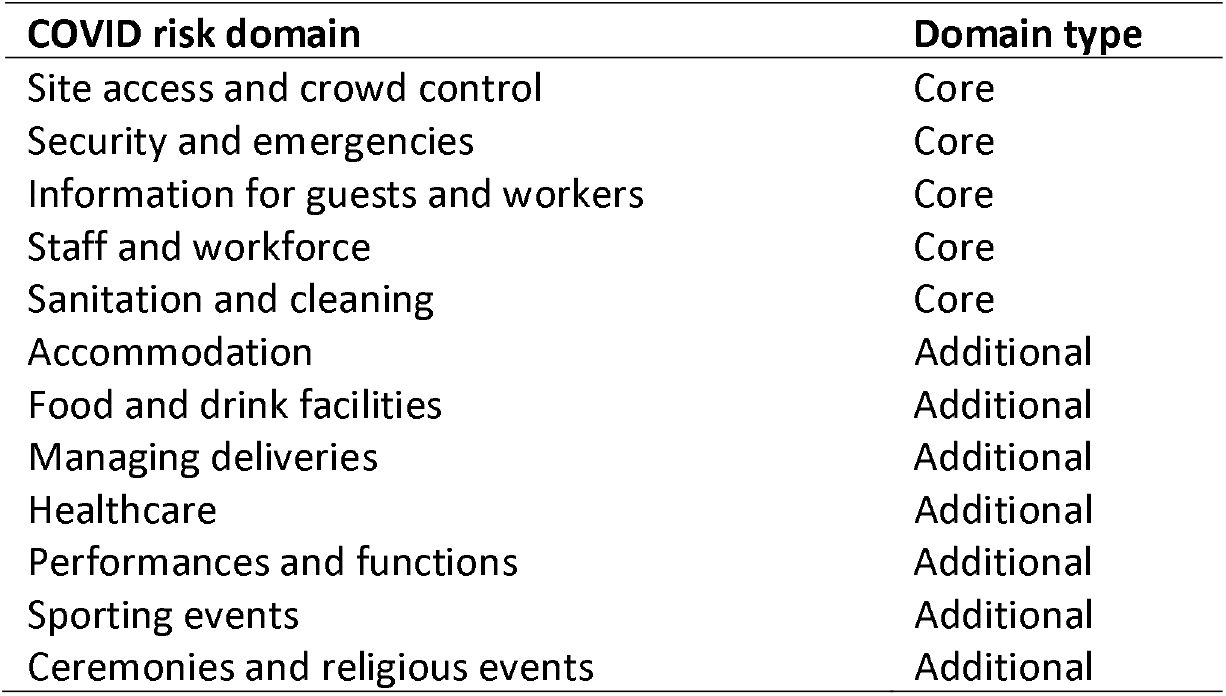
COVID risk domains listed in the C-SET framework, by type.

Note that there may be overlap in guidance (e.g. the visitor economy guidance also applies to the performing arts), and the scope of businesses to which the guidance applies may be greater than the name implies (e.g. performing arts guidance also relates to speakers at conferences or events). For further details on the scope of each guidance, please consult the relevant guidance.

Although not linked to specific articles of national guidance, other considerations should be made to the context of the region in which the event is taking place. These have been agreed based on expert views and local consensus. They fall into two broad categories: considerations relating to the event specifically, but not mentioned in the national guidance, and considerations of the local and national context in which these events are taking place.

Event-related considerations are those recommended by colleagues responsible for either licensing the event and or ensuring that it is COVID-secure, and have been implemented locally in Suffolk. These considerations include:

- Asking event organisers to provide the region of origin of attendees and staff (including contractors) so that risk of widespread transmission or cross-region spread could be assessed by event reviewers
- Ensuring that no one attends from areas of medium or high COVID alert levels from the three-tiered national model of restrictions published on GOV.UK^16^
- Assessing the risks posed to protected groups
- Ensuring that events have a clear policy outlining refund procedures in case of COVID-related issues (to facilitate attendee adherence to public health guidance e.g. staying home if they have symptoms consistent with COVID-19)

Considerations which relate to the local and national context are intended to account for the fact that these events do not take place in isolation, and that although an event may itself be COVID-secure, there may be external considerations which may impact on this. These considerations include:

- Gathering disease prevalence and transmission rates in the area
- Multiple ongoing outbreaks in the area in different settings (e.g. schools, workplaces)
- Assessing the cumulative risk of multiple large events taking place within a short space of time
- Current and predicted trend in number of COVID cases and impact on the local and national NHS

### Structure of the checklist

The checklist is derived from the framework; in addition to questions around each key domain, it collects additional information on the event characteristics (“event profile” section of the checklist). The checklist process and who fills it hasn’t been explicitly prescribed by the authors; however, the suggestion is that the event organisers provide evidence against each checklist question, following which a “reviewer” (e.g. a local SAG member) would rate all the evidence provided (a red-amber-green (RAG) rating system is built into the dropdowns in the checklist).

A quick summary of the event is collected on the “event profile” section at the beginning of the checklist. It includes information on the nature and location of the event, anticipated numbers of attendees, age range and composition of attendees (including staff and contractors). Requests are made to quantify available toilet facilities where possible, to allow mitigations to account for levels of use and risks of interaction and spread. These are likely to impact upon the mitigations required (e.g. more stringent requirements for international events, or those with vulnerable attendees).

The event profile section also asks whether certain facilities are available at the event (e.g. accommodation, food/retail facilities); these are screening questions, and answers to these will direct checklist users to complete the appropriate section of the checklist. Section 1 is comprised of the common COVID risk domains outlined in the framework, and should be completed for all events; later sections (sections 2-7) may only be relevant to certain events as indicated in the earlier screening questions.

A summary of areas of concern is automatically generated in the “summary” section of the checklist. Domain- and subdomain-specific recommendations can then be made by referring to the framework document to enable organisers to make positive changes to their events.

To help in identifying areas of weakness, this summary is provided per COVID risk domain using a RAG rating system. A second breakdown per individual sub-domain is also available for more detailed examination. This is intended to allow development of specific, actionable recommendations to event organisers.

The checklist summary does not include in-built thresholds for an overall rating of events, as these are likely to vary based upon the type of event and the body undertaking the analysis. A suggested rating system is a three-tier rating similar to the RAG system. For example:

- No concerns – event mitigations are adequate. Event can proceed as planned.
- Minor concerns – event can proceed with minor modifications which, taken together, are unlikely to affect the administration or running of the event.
- Major concerns – event requires major modifications to be COVID-secure; these may require significant organisational restructuring and may result in a materially different event. Depending on timeframes, it may be appropriate to consider delaying or cancelling the event.

## Discussion

### What is already known on the topic

Additional considerations are required to ensure that events taking place while COVID-19 is in circulation to minimise the risk of onward transmission to attendees and staff. Peers in this area, most notably the WHO Novel Coronavirus-19 Mass Gatherings Expert Group^8^, have proposed basic guidelines. However, they do not provide specific mitigations, nor do they provide a mechanism by which to provide summary recommendations. Professional experience suggests that overall approaches to identifying risk areas vary dramatically by sector, resource and event type.

### What this study adds

To our knowledge, C-SET is the first toolkit created to assist large events taking place in the COVID-19 era offering a structure for mitigations, and is the most comprehensive toolkit to date. The methods used to develop this toolkit are similar to those used elsewhere: others have incorporated pilots^17^ and expert consultation^18,19^, and the authors chose to include both to maximise usability and functionality of the toolkit.

C-SET offers a robust, structured approach to identifying areas of concern and potential mitigations, and collates and synthesises current national UK guidance. A flexible, modular design allows it to provide high-level input across all types of event, and allows it to be modified to suit the needs of individual organisations. The agnostic design makes it suitable for a variety of users, including event organisers in need of a structure, or bodies whose role is to appraise such events. A user-friendly checklist provides a semi-quantitative summary output, and in conjunction with the detailed links to guidance in the framework, allows development of clear, actionable recommendations to tangibly improve the quality of events.

The authors believe that C-SET fills a void that currently exists in assessing large gatherings in the COVID era. Our early experience in Suffolk indicates use of some of the principles and contents of the framework has led one event organiser to radically transform their initial plans. This has reduced the risk of COVID transmission greatly, making the event COVID secure and at the same time supporting the local economy (Personal communication – Chair of Suffolk SAGs). With the holiday season fast approaching, many more such events might be planned across the country. C-SET will be a timely tool to make these events as COVID secure as possible.

### Limitations of the study

In order to remain user-friendly but comprehensive, some of the guidance is generic in nature: depending upon the specific scope and context of the event, certain sections may not be relevant (e.g. if there is no overnight accommodation being provided as part of the event). Furthermore, there may be subtle differences between modifications required in specific circumstances; this toolkit tends to utilise high-level groupings, with specific additional guidance provided in the corresponding section of the framework. For example, while camp-sites may have different requirements to dormitories, common themes are listed in C-SET, and this is advised in framework.

Suggested mitigations outlined within C-SET are not exhaustive. Other mitigations may be appropriate, according to the specifics of the event; however, it is anticipated that those using this toolkit will have subject expertise sufficient to make these decisions.

In addition, this toolkit does not account for activities associated with preparation for an event, such as rehearsals ahead of performing arts events. These considerations, while important, are likely to be very different from one event to another, and capturing the full breadth of these would negatively impact usability. However, the principles outlined in C-SET may be used to inform future work in this area.

National guidance is changing in line with the evolving COVID-19 situation. Whilst several principles are likely to apply throughout the pandemic (e.g. hand-washing, social distancing, enhanced cleaning), the specifics of these may to change. Maintaining the toolkit is critical to its ongoing relevance and applicability, but it is the authors’ view that subject matter experts who will be using this on a daily basis will take ownership and keep it up to date with support from Public Health colleagues in their respective areas.

In order to remain as evidence-based as possible, this framework limits the number of additional local considerations which have been included after consultation with experts. C-SET may be adapted as necessary to reflect local and regional policies, and expert opinion.

## Supporting information

Appendix 1: C-SET framework

Appendix 2: C-SET checklist

Appendix 3: C-SET user manual

## Data Availability

All relevant source material is available via hyperlink in supplementary material.

## Acknowledgements

The authors are grateful to John Grayling, V Johnston, Gavin Oakenfull and Mark Skillin (chairs and members of the Suffolk SAGs) for their expert input during the consultation, and to Stuart Keeble, Director of Public Health, Suffolk County Council for his support and encouragement.

