## Appendix 1: C-SET framework for "The COVID-19 Suffolk Events Toolkit (C-SET): A structured approach to conducting COVID-secure events"

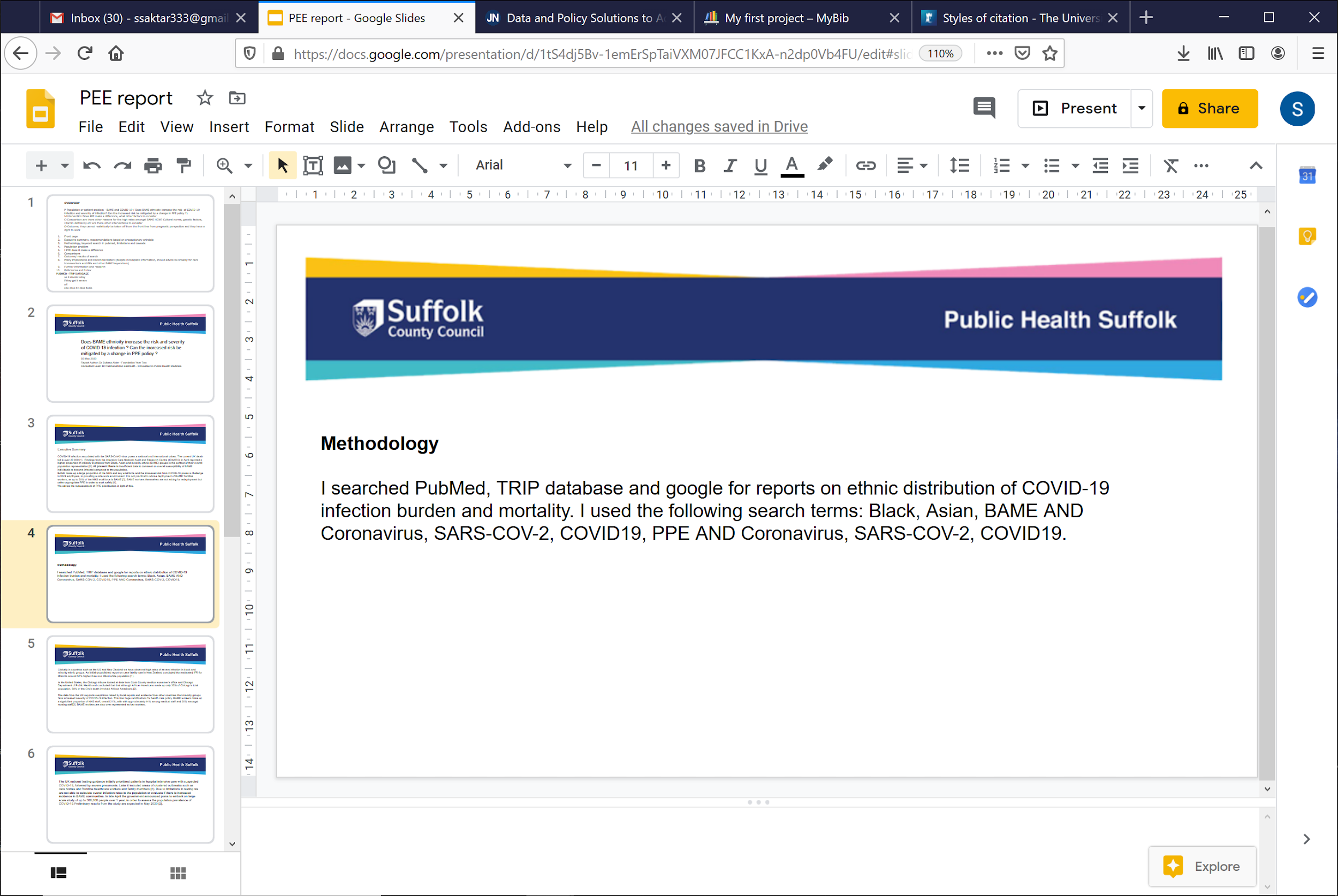

**The COVID-19 Suffolk Events Toolkit (C-SET) framework: facilitating COVID-secure gatherings**

Public Health Suffolk – Suffolk County Council

21 October 2020

**Authors**

Dr Avirup Chowdhury^1^, Terezija Zermanos^2^, Dr Xinming Yu, Dr Padmanabhan Badrinath^3^

^1^Specialty Registrar in Public Health Medicine, Public Health Suffolk, Suffolk County Council, Ipswich

^2^Health and Care Programme Manager, Public Health Suffolk, Suffolk County Council, Ipswich

^3^Foundation Doctor in Public Health Medicine, Public Health Suffolk, Suffolk County Council, Ipswich

^3^Consultant in Public Health Medicine, Public Health Suffolk, Suffolk County Council, Ipswich

### 1 Introduction

Since the beginning of the COVID-19 global pandemic, the negative impact of COVID-19 transmission (in the form of increased morbidity and mortality) has been well-established. However, there are also negative consequences as a result of prolonged economic depression, as well as health consequences of prolonged lockdown (e.g. worsening mental health outcomes). This has given rise to a tension between the economic and health advantages of opening the economy and the negative consequences (in particular, an increase in COVID-19 transmission and mortality). Large gatherings are a key manifestation of this dilemma: they often bring significant economic activity to a local region, but risks nonetheless exist when potentially thousands of people living in areas with differing COVID-19 prevalence travel long distances to attend a single event.

Creation of so-called COVID-secure events may allow a positive trade-off between these opposing interests, and may allow leveraging of the positive effects of large events while minimising the negative COVID-19 health risks to the wider community. Given the broad variation in size, scope and nature of these large gatherings, it is not unexpected that there has been a similar variation in the mitigations that have been implemented by organisations, and the types of documentation used to report these to relevant bodies.

The COVID-19 Suffolk Events Toolkit (C-SET) has been created to facilitate a systematic appraisal of best practices to minimise risks arising as a consequence of large events during the COVID-19 pandemic, drawing upon national guidance to define key risk domains. It has not been designed with a single user-group in mind, and may provide a guide for organisations planning COVID-secure events, or aid statutory bodies in appraising events and suggesting mitigations. It provides general principles which are applicable to all events, with additional sections for specific risk areas (e.g. accommodation, retail) which may not apply universally. This is discussed further in section 1.1.2.

Please note that the guidance below is generic in nature: depending upon the specific scope and context of the event, certain sections may not be relevant (e.g. if there is no overnight accommodation being provided as part of the event). In addition, while suggested mitigations are outlined below (and are according to national guidance), these should not be considered exhaustive. Other mitigations may be appropriate, according to the specifics of the event.

C-SET encompasses three key documents:

- Framework
- Checklist
- User manual

This framework outlines key COVID domains and considerations within these areas including suggested mitigations to ensure that events are COVID-secure with links to appropriate national guidance. It also provides the rationale which underpins C-SET. National guidance links are correct at the time of writing but may be subject to change. However, the general principles may continue to apply (e.g. handwashing, social distancing).

A checklist has been developed on the basis of this framework. It can be used to determine whether an event provides sufficient mitigations across a comprehensive set of COVID risk domains, condensing the key risk domains and subdomains in the framework into questions across various sections.

The user manual provides a practical guide for those using the checklist, and is designed to accompany it.

#### 1.1 Gathering evidence

This section briefly outlines the information gathered using the C-SET checklist. These are generic suggestions, and individual organisations may find utility in tailoring this toolkit to their needs.

##### 1.1.1 Event profile

A quick summary of the event is collected on the first page of the checklist. It includes:

- the nature of the event (e.g. music festival)
- date(s) of event
- location of event, including whether it may be cross-boundary with other LAs
- event organiser(s)

It should also include a brief outline of the profile of attendees (age, sex, family vs non-family groups, place of origin i.e. national/international), sleeping (e.g. camping, dormitories) and eating arrangements (e.g. self-catering, food stalls) as these will influence the scope of the risk assessment and subsequent recommendations. This is part of the first section of the accompanying checklist.

Requests are made to quantify available facilities and staff where possible, to allow mitigations to account for levels of use and risks of interaction and spread.

##### 1.1.2 COVID risk domains

Where possible, multiple links to guidance have been provided in this framework. A shortened title and section number have been provided in each instance. Please note that there may be overlap in guidance (e.g. the visitor economy guidance also applies to the performing arts), and the scope of businesses to which the guidance applies may be greater than the name implies (e.g. performing arts guidance also relates to speakers at conferences or events). For further details on the scope of each guidance, please follow the relevant link.

In addition, COVID risk domains in sections 2-6 are likely to apply to all large gatherings, while later sections may only be appropriate to certain events (e.g. those with on-site medical facilities); the final three risk domains apply only to specific high-risk events (performing arts, sporting events and ceremonies and religious events). These essential risk domains are covered in section 1 of the checklist, with optional domains in later sections. Further detail is available in the checklist user manual.

##### 1.1.3 Providing recommendations

In conjunction to COVID risk domains outlined in this document, there are additional considerations which should be taken into account when providing recommendations to events. These are covered in more detail in Section 13.

When completing the checklist, a summary of areas of concern is automatically generated in the summary tab. Domain- and subdomain- specific recommendations can then be made by referring to this framework, to enable organisers to make positive changes to their events.

For bodies assessing COVID-security using this framework, it may be helpful to provide an overall recommendation to organisers regarding the event as a whole. This could take several forms, including a three-tier rating. For example:

- No concerns – event mitigations are adequate. Event can proceed as planned.
- Minor concerns – event can proceed with minor modifications which, taken together, are unlikely to affect the administration or running of the event.
- Major concerns – event requires major modifications to be COVID-secure; these may require significant organisational restructuring and may result in a materially different event. Depending on timeframes, it may be appropriate to consider delaying or cancelling the event.

### 2 COVID risk domain: Site access and crowd control

#### 2.1 Site capacity

| **Key considerations** | **Potential mitigations** | **Suggested questions** | **Relevant guidance** |
| --- | --- | --- | --- |
| Capacity calculations | Reducing site, premises or venue capacity and limiting ticket sales to a volume which ensures social distancing can be maintained. This will vary depending on layout or usage. This will require taking into account the total floorspace as well as pinch points and busy areas. | What is the usual (pre-COVID) maximum capacity of the site, and what is the maximum capacity to remain COVID-secure? | [Hotels and guest accommodation 2.2](https://www.gov.uk/guidance/working-safely-during-coronavirus-covid-19/hotels-and-other-guest-accommodation#section-2-2)  [Performing arts 3.2](https://www.gov.uk/guidance/working-safely-during-coronavirus-covid-19/performing-arts#arts-2-1)  [Performing arts 3.3](https://www.gov.uk/guidance/working-safely-during-coronavirus-covid-19/performing-arts#arts-3-3)  [Places of worship](https://www.gov.uk/government/publications/covid-19-guidance-for-the-safe-use-of-places-of-worship-during-the-pandemic-from-4-july/covid-19-guidance-for-the-safe-use-of-places-of-worship-during-the-pandemic-from-4-july)  [Restaurants, pubs, bars and takeaway services 2.1](https://www.gov.uk/guidance/working-safely-during-coronavirus-covid-19/restaurants-offering-takeaway-or-delivery#takeaways-2-1)  [Return of sport](https://www.gov.uk/government/publications/coronavirus-covid-19-guidance-on-phased-return-of-sport-and-recreation/guidance-for-the-public-on-the-phased-return-of-outdoor-sport-and-recreation)  [Visitor economy 2.1](https://www.gov.uk/guidance/working-safely-during-coronavirus-covid-19/the-visitor-economy#shops-4-1) |
|  | Limiting the number of people in the venue or on the premises, overall and in any particular congestion areas, for example, doorways between outside and inside spaces. This will vary depending on layout or usage. This will require taking into account the total floorspace as well as pinch points and busy areas. | What is the usual (pre-COVID) maximum capacity of the site, and what is the maximum capacity to remain COVID-secure? | [Performing arts 3.1](https://www.gov.uk/guidance/working-safely-during-coronavirus-covid-19/performing-arts#arts-3-1)  [Restaurants, pubs, bars and takeaway services 2.1](https://www.gov.uk/guidance/working-safely-during-coronavirus-covid-19/restaurants-offering-takeaway-or-delivery#takeaways-2-1)  [Return of sport](https://www.gov.uk/government/publications/coronavirus-covid-19-guidance-on-phased-return-of-sport-and-recreation/guidance-for-the-public-on-the-phased-return-of-outdoor-sport-and-recreation)  [Visitor economy 2.1](https://www.gov.uk/guidance/working-safely-during-coronavirus-covid-19/the-visitor-economy#shops-4-1) |
|  | Using the Crowd Density Standard for indoor events and shows: at a capacity allowing for compliance with social distancing of 2m, or 1m with mitigation (approximately equivalent to a density of 10m^2^ per person). | What density standard have you used (stating number of m^2^ per person & equivalent social distance)? | [Visitor economy 2.2](https://www.gov.uk/guidance/working-safely-during-coronavirus-covid-19/the-visitor-economy#shops-4-2) |
|  | Using, for seated accommodation in sports grounds, a 2.5m diameter circle, and for standing areas or concourses, for calculation purposes, a 2.6m square (6.8 m^2^ per person). | What density standard have you used (stating number of m^2^ per person & equivalent social distance)? | [SG02 2.2](https://sgsa.org.uk/wp-content/uploads/2020/08/SG02-Planning-for-Social-Distancing-at-Sports-Grounds.pdf) |
| Risk of larger groups forming | Local authorities should avoid issuing licenses for events that could lead to larger gatherings forming and provide advice to businesses on how to manage events of this type. | What is the risk of larger groups forming? | [Performing arts 3.3](https://www.gov.uk/guidance/working-safely-during-coronavirus-covid-19/performing-arts#arts-3-3)  [Return of sport](https://www.gov.uk/government/publications/coronavirus-covid-19-guidance-on-phased-return-of-sport-and-recreation/guidance-for-the-public-on-the-phased-return-of-outdoor-sport-and-recreation) |

#### 2.2 Transport to site

| **Key considerations** | **Potential mitigations** | **Suggested questions** | **Relevant guidance** |
| --- | --- | --- | --- |
| Transport choices | Support attendees and staff to walk and cycle if they can, avoiding using public transport. | How have you supported attendees to walk and cycle, and avoid public transport? | [Performing arts 3.5](https://www.gov.uk/guidance/working-safely-during-coronavirus-covid-19/performing-arts#arts-3-5)  [Performing arts 4.17](https://www.gov.uk/guidance/working-safely-during-coronavirus-covid-19/performing-arts#arts-4-17)  [Restaurants, pubs, bars and takeaway services 2.1](https://www.gov.uk/guidance/working-safely-during-coronavirus-covid-19/restaurants-offering-takeaway-or-delivery#takeaways-2-1)  [Visitor economy 2.2](https://www.gov.uk/guidance/working-safely-during-coronavirus-covid-19/the-visitor-economy#shops-4-2) |
|  | Limiting passengers in shared vehicles, e.g. by leaving seats empty. | If providing shared vehicles, how are you limiting passenger numbers? | [Performing arts 3.5](https://www.gov.uk/guidance/working-safely-during-coronavirus-covid-19/performing-arts#arts-3-5)  [Visitor economy 2.2](https://www.gov.uk/guidance/working-safely-during-coronavirus-covid-19/the-visitor-economy#shops-4-2) |
| Access from drop-off to entrance | Arranging one-way travel routes between transport hubs and venues. | Where are the nearest transport hubs, and how are you arranging one-way travel routes between transport hubs and venues? | [Hotels and guest accommodation 2.2](https://www.gov.uk/guidance/working-safely-during-coronavirus-covid-19/hotels-and-other-guest-accommodation#section-2-2)  [Performing arts 3.3](https://www.gov.uk/guidance/working-safely-during-coronavirus-covid-19/performing-arts#arts-3-3)  [Places of worship](https://www.gov.uk/government/publications/covid-19-guidance-for-the-safe-use-of-places-of-worship-during-the-pandemic-from-4-july/covid-19-guidance-for-the-safe-use-of-places-of-worship-during-the-pandemic-from-4-july)  [Visitor economy 2.1](https://www.gov.uk/guidance/working-safely-during-coronavirus-covid-19/the-visitor-economy#shops-4-1)  [Visitor economy 2.2](https://www.gov.uk/guidance/working-safely-during-coronavirus-covid-19/the-visitor-economy#shops-4-2) |
|  | Advising patrons to avoid particular forms of transport or routes and to avoid crowded areas when in transit to the venue. | How are you advising patrons to avoid particular forms of transport or routes and to avoid crowded areas when in transit to the venue? | [Performing arts 3.3](https://www.gov.uk/guidance/working-safely-during-coronavirus-covid-19/performing-arts#arts-3-3)  [Hotels and guest accommodation 2.2](https://www.gov.uk/guidance/working-safely-during-coronavirus-covid-19/hotels-and-other-guest-accommodation#section-2-2)  [Visitor economy 2.1](https://www.gov.uk/guidance/working-safely-during-coronavirus-covid-19/the-visitor-economy#shops-4-1) |

#### 2.3 Car parking

| **Key considerations** | **Potential mitigations** | **Suggested questions** | **Relevant guidance** |
| --- | --- | --- | --- |
| Capacity calculations | Reducing site, premises or venue capacity and limiting ticket sales to a volume which ensures social distancing can be maintained. | What is the maximum (pre-COVID) capacity of the car park, and what is the maximum capacity to remain COVID-secure? | [Performing arts 3.2](https://www.gov.uk/guidance/working-safely-during-coronavirus-covid-19/performing-arts#arts-2-1)  [Places of worship 5](https://www.gov.uk/government/publications/covid-19-guidance-for-the-safe-use-of-places-of-worship-during-the-pandemic-from-4-july/covid-19-guidance-for-the-safe-use-of-places-of-worship-during-the-pandemic-from-4-july)  [Visitor economy 2.1](https://www.gov.uk/guidance/working-safely-during-coronavirus-covid-19/the-visitor-economy#shops-4-1) |
|  | Planning car parking to allow sufficient spacing for the social distancing of occupants. | What modifications have you made to allow sufficient spacing for the social distancing of occupants? | [Performing arts 3.4](https://www.gov.uk/guidance/working-safely-during-coronavirus-covid-19/performing-arts#arts-3-4)  [Places of worship 5](https://www.gov.uk/government/publications/covid-19-guidance-for-the-safe-use-of-places-of-worship-during-the-pandemic-from-4-july/covid-19-guidance-for-the-safe-use-of-places-of-worship-during-the-pandemic-from-4-july)  [Visitor economy 2.2](https://www.gov.uk/guidance/working-safely-during-coronavirus-covid-19/the-visitor-economy#shops-4-2) |

#### 2.4 Ingress and egress

| **Key considerations** | **Potential mitigations** | **Suggested questions** | **Relevant guidance** |
| --- | --- | --- | --- |
| One-way systems | Using markings and introducing an accessible one-way flow at entry and exit points. | Have you introduced a one-way system, and if so, what markings are you using to signpost this? Have you utilised these at entry and exit points? | [Performing arts 3.5](https://www.gov.uk/guidance/working-safely-during-coronavirus-covid-19/performing-arts#arts-3-5)  [Performing arts 3.11](https://www.gov.uk/guidance/working-safely-during-coronavirus-covid-19/performing-arts#arts-3-11)  [Places of worship 5](https://www.gov.uk/government/publications/covid-19-guidance-for-the-safe-use-of-places-of-worship-during-the-pandemic-from-4-july/covid-19-guidance-for-the-safe-use-of-places-of-worship-during-the-pandemic-from-4-july)  [Visitor economy 2.1](https://www.gov.uk/guidance/working-safely-during-coronavirus-covid-19/the-visitor-economy#shops-4-1)  [Visitor economy 2.2](https://www.gov.uk/guidance/working-safely-during-coronavirus-covid-19/the-visitor-economy#shops-4-2) |
| Queue and crowd control | Staggering entry and departure times. | Have you staggered entry and departure times? If so, please provide details. | [Hotels and guest accommodation 2.2](https://www.gov.uk/guidance/working-safely-during-coronavirus-covid-19/hotels-and-other-guest-accommodation#section-2-2)  [Performing arts 3.3](https://www.gov.uk/guidance/working-safely-during-coronavirus-covid-19/performing-arts#arts-3-3)  [Performing arts 3.5](https://www.gov.uk/guidance/working-safely-during-coronavirus-covid-19/performing-arts#arts-3-5)  [Places of worship](https://www.gov.uk/government/publications/covid-19-guidance-for-the-safe-use-of-places-of-worship-during-the-pandemic-from-4-july/covid-19-guidance-for-the-safe-use-of-places-of-worship-during-the-pandemic-from-4-july)  [Visitor economy 2.1](https://www.gov.uk/guidance/working-safely-during-coronavirus-covid-19/the-visitor-economy#shops-4-1)  [Visitor economy 2.2](https://www.gov.uk/guidance/working-safely-during-coronavirus-covid-19/the-visitor-economy#shops-4-2) |
|  | Staggering entry times with other venues and taking steps to avoid queues building up in surrounding areas. | Have you staggered entry and departure times with other venues? If so, please provide details. | [Hotels and guest accommodation 2.2](https://www.gov.uk/guidance/working-safely-during-coronavirus-covid-19/hotels-and-other-guest-accommodation#section-2-2)  [Performing arts 3.3](https://www.gov.uk/guidance/working-safely-during-coronavirus-covid-19/performing-arts#arts-3-3)  [Performing arts 3.5](https://www.gov.uk/guidance/working-safely-during-coronavirus-covid-19/performing-arts#arts-3-5)  [Places of worship](https://www.gov.uk/government/publications/covid-19-guidance-for-the-safe-use-of-places-of-worship-during-the-pandemic-from-4-july/covid-19-guidance-for-the-safe-use-of-places-of-worship-during-the-pandemic-from-4-july)  [Visitor economy 2.1](https://www.gov.uk/guidance/working-safely-during-coronavirus-covid-19/the-visitor-economy#shops-4-1)  [Visitor economy 2.2](https://www.gov.uk/guidance/working-safely-during-coronavirus-covid-19/the-visitor-economy#shops-4-2) |
|  | Calculating rates of flow, based on references (see link), to ensure social distancing can be maintained throughout the venue, and on ingress and egress. | Please provide details on the standard used to calculate rates of person flow through the venue. | [SG02 5](https://sgsa.org.uk/wp-content/uploads/2020/08/SG02-Planning-for-Social-Distancing-at-Sports-Grounds.pdf)  [SG02 6](https://sgsa.org.uk/wp-content/uploads/2020/08/SG02-Planning-for-Social-Distancing-at-Sports-Grounds.pdf)  [SG02 9](https://sgsa.org.uk/wp-content/uploads/2020/08/SG02-Planning-for-Social-Distancing-at-Sports-Grounds.pdf) |
| Cloakrooms | Closing cloakrooms wherever possible. | Have you opened cloakrooms in the venue? | [Performing arts 3.5](https://www.gov.uk/guidance/working-safely-during-coronavirus-covid-19/performing-arts#arts-3-5) |
| Equality of access | Considering the particular needs of disabled audiences when making adjustments to venues or premises to ensure no disadvantage. | How have you ensured that disabled participants will not be disadvantaged by changes to ingress and egress? | [Performing arts 3.2](https://www.gov.uk/guidance/working-safely-during-coronavirus-covid-19/performing-arts#arts-2-1)  [Places of worship](https://www.gov.uk/government/publications/covid-19-guidance-for-the-safe-use-of-places-of-worship-during-the-pandemic-from-4-july/covid-19-guidance-for-the-safe-use-of-places-of-worship-during-the-pandemic-from-4-july)  [Restaurants, pubs, bars and takeaway services 2.1](https://www.gov.uk/guidance/working-safely-during-coronavirus-covid-19/restaurants-offering-takeaway-or-delivery#takeaways-2-1)  [Visitor economy 2.1](https://www.gov.uk/guidance/working-safely-during-coronavirus-covid-19/the-visitor-economy#shops-4-1) |
|  | Ensure any changes to entries, exit and queue management take into account reasonable adjustments for those who need them. | How have you ensured that disabled participants will not be disadvantaged by changes to ingress and egress? | [Performing arts 3.5](https://www.gov.uk/guidance/working-safely-during-coronavirus-covid-19/performing-arts#arts-3-5)  [Performing arts 3.11](https://www.gov.uk/guidance/working-safely-during-coronavirus-covid-19/performing-arts#arts-3-11)  [Places of worship](https://www.gov.uk/government/publications/covid-19-guidance-for-the-safe-use-of-places-of-worship-during-the-pandemic-from-4-july/covid-19-guidance-for-the-safe-use-of-places-of-worship-during-the-pandemic-from-4-july)  [Restaurants, pubs, bars and takeaway services 2.1](https://www.gov.uk/guidance/working-safely-during-coronavirus-covid-19/restaurants-offering-takeaway-or-delivery#takeaways-2-1)  [Visitor economy 2.1](https://www.gov.uk/guidance/working-safely-during-coronavirus-covid-19/the-visitor-economy#shops-4-1) |
|  | Considering how social distancing markers can be made as accessible as reasonably practicable. | How have you made social distancing markers accessible to all attendees? | [Performing arts 3.11](https://www.gov.uk/guidance/working-safely-during-coronavirus-covid-19/performing-arts#arts-3-11)  [Visitor economy 2.1](https://www.gov.uk/guidance/working-safely-during-coronavirus-covid-19/the-visitor-economy#shops-4-1) |
| Locations | Having more entry points in larger premises or venues. | Have you increased the number of entry points in your venue? Please provide details. | [Performing arts 3.5](https://www.gov.uk/guidance/working-safely-during-coronavirus-covid-19/performing-arts#arts-3-5)  [Places of worship 5](https://www.gov.uk/government/publications/covid-19-guidance-for-the-safe-use-of-places-of-worship-during-the-pandemic-from-4-july/covid-19-guidance-for-the-safe-use-of-places-of-worship-during-the-pandemic-from-4-july) |
|  | Using e.g. auditorium fire exits as the standard so that guests are not required to pass each other when entering and exiting spaces. | How have you ensured that guests are not required to pass one another when entering and exiting spaces? | [Performing arts 3.11](https://www.gov.uk/guidance/working-safely-during-coronavirus-covid-19/performing-arts#arts-3-11)  [Places of worship 5](https://www.gov.uk/government/publications/covid-19-guidance-for-the-safe-use-of-places-of-worship-during-the-pandemic-from-4-july/covid-19-guidance-for-the-safe-use-of-places-of-worship-during-the-pandemic-from-4-july) |
| Entry procedures | Providing handwashing facilities (or hand sanitiser) at entry and exit points and encouraging their use. | Have you provided handwashing facilities (or hand sanitiser) at entry and exit points? | [Performing arts 3.5](https://www.gov.uk/guidance/working-safely-during-coronavirus-covid-19/performing-arts#arts-3-5)  [Performing arts 3.11](https://www.gov.uk/guidance/working-safely-during-coronavirus-covid-19/performing-arts#arts-3-11)  [Restaurants, pubs, bars and takeaway services 4.1](https://www.gov.uk/guidance/working-safely-during-coronavirus-covid-19/restaurants-offering-takeaway-or-delivery#takeaways-4-1)  [Food businesses](https://www.gov.uk/government/publications/covid-19-guidance-for-food-businesses/guidance-for-food-businesses-on-coronavirus-covid-19)  [Visitor economy 2.2](https://www.gov.uk/guidance/working-safely-during-coronavirus-covid-19/the-visitor-economy#shops-4-2) |
|  | Providing alternatives to touch-based security devices such as keypads. | If appropriate, have you provided alternatives to touch-based security devices such as keypads? | [Performing arts 3.5](https://www.gov.uk/guidance/working-safely-during-coronavirus-covid-19/performing-arts#arts-3-5) |
|  | Defining process alternatives for entry/exit points where appropriate. | Have you defined process alternatives for entry/exit points where appropriate? | [Performing arts 3.5](https://www.gov.uk/guidance/working-safely-during-coronavirus-covid-19/performing-arts#arts-3-5) |
| Emergency egress | In planning the event site, consideration needs to be given to evacuating attendees in case of emergency in such a way as to maintain social distancing, where possible. | Please outline how you will ensure evacuation of attendees in emergency while maintaining social distancing. | [Events Industry Forum 4.4](https://www.eventindustrynews.com/wp-content/uploads/2020/07/EIfDCMS-COVID-19-Working-Safely-9-July-2020.pdf) |

#### 2.5 Crowd control

| **Key considerations** | **Potential mitigations** | **Suggested questions** | **Relevant guidance** |
| --- | --- | --- | --- |
| Queueing | Using space outside the site, premises or venue for queuing where available and safe. | Have you used space outside the venue for queueing? If not, why not? | [Performing arts 3.11](https://www.gov.uk/guidance/working-safely-during-coronavirus-covid-19/performing-arts#arts-3-11)  [Visitor economy 2.1](https://www.gov.uk/guidance/working-safely-during-coronavirus-covid-19/the-visitor-economy#shops-4-1)  [Visitor economy 2.2](https://www.gov.uk/guidance/working-safely-during-coronavirus-covid-19/the-visitor-economy#shops-4-2) |
|  | Reducing instances where people might be required to queue. | How have you reduced instances where people might be required to queue? | [Food businesses](https://www.gov.uk/government/publications/covid-19-guidance-for-food-businesses/guidance-for-food-businesses-on-coronavirus-covid-19)  [Performing arts 3.11](https://www.gov.uk/guidance/working-safely-during-coronavirus-covid-19/performing-arts#arts-3-11)  [Places of worship 5](https://www.gov.uk/government/publications/covid-19-guidance-for-the-safe-use-of-places-of-worship-during-the-pandemic-from-4-july/covid-19-guidance-for-the-safe-use-of-places-of-worship-during-the-pandemic-from-4-july)  [Restaurants, pubs, bars and takeaway services 2.1](https://www.gov.uk/guidance/working-safely-during-coronavirus-covid-19/restaurants-offering-takeaway-or-delivery#takeaways-2-1)  [Visitor economy 2.1](https://www.gov.uk/guidance/working-safely-during-coronavirus-covid-19/the-visitor-economy#shops-4-1) |
|  | Designating staff to manage queues and regulate guest access. | Have you designated staff to manage queues? | [Performing arts 3.11](https://www.gov.uk/guidance/working-safely-during-coronavirus-covid-19/performing-arts#arts-3-11)  [Places of worship 5](https://www.gov.uk/government/publications/covid-19-guidance-for-the-safe-use-of-places-of-worship-during-the-pandemic-from-4-july/covid-19-guidance-for-the-safe-use-of-places-of-worship-during-the-pandemic-from-4-july)  [Visitor economy 2.1](https://www.gov.uk/guidance/working-safely-during-coronavirus-covid-19/the-visitor-economy#shops-4-1) |
|  | Helping visitors and staff maintain social distancing by placing clearly visible markers along the ground, floor or walls. | What markings are you using to signpost social distancing? | [Food businesses](https://www.gov.uk/government/publications/covid-19-guidance-for-food-businesses/guidance-for-food-businesses-on-coronavirus-covid-19)  [Performing arts 3.11](https://www.gov.uk/guidance/working-safely-during-coronavirus-covid-19/performing-arts#arts-3-11)  [Places of worship 5](https://www.gov.uk/government/publications/covid-19-guidance-for-the-safe-use-of-places-of-worship-during-the-pandemic-from-4-july/covid-19-guidance-for-the-safe-use-of-places-of-worship-during-the-pandemic-from-4-july)  [Restaurants, pubs, bars and takeaway services 4.3](https://www.gov.uk/guidance/working-safely-during-coronavirus-covid-19/restaurants-offering-takeaway-or-delivery#takeaways-4-3)  [Visitor economy 2.2](https://www.gov.uk/guidance/working-safely-during-coronavirus-covid-19/the-visitor-economy#shops-4-2) |
|  | Having clearly designated positions from which employees can provide assistance to customers whilst maintaining social distance. | Are you designating employees to provide assistance to customers? If so, how are you ensuring they socially distance? | [Visitor economy 2.1](https://www.gov.uk/guidance/working-safely-during-coronavirus-covid-19/the-visitor-economy#shops-4-1) |
|  | Managing outside queues to ensure they do not cause a risk to individuals or other businesses. | How are you ensuring that queues outside do not pose a risk to individuals or other businesses? | [Hotels and guest accommodation 2.2](https://www.gov.uk/guidance/working-safely-during-coronavirus-covid-19/hotels-and-other-guest-accommodation#section-2-2)  [Places of worship](https://www.gov.uk/government/publications/covid-19-guidance-for-the-safe-use-of-places-of-worship-during-the-pandemic-from-4-july/covid-19-guidance-for-the-safe-use-of-places-of-worship-during-the-pandemic-from-4-july)  [Visitor economy 2.2](https://www.gov.uk/guidance/working-safely-during-coronavirus-covid-19/the-visitor-economy#shops-4-2) |
| Reducing pinch points | Introducing more one-way flow through buildings. | Have you introduced a one-way system through the building? | [Performing arts 3.6](https://www.gov.uk/guidance/working-safely-during-coronavirus-covid-19/performing-arts#arts-3-6)  [Places of worship 5](https://www.gov.uk/government/publications/covid-19-guidance-for-the-safe-use-of-places-of-worship-during-the-pandemic-from-4-july/covid-19-guidance-for-the-safe-use-of-places-of-worship-during-the-pandemic-from-4-july)  [Restaurants, pubs, bars and takeaway services 4.4](https://www.gov.uk/guidance/working-safely-during-coronavirus-covid-19/restaurants-offering-takeaway-or-delivery#takeaways-4-4)  [Visitor economy 2.1](https://www.gov.uk/guidance/working-safely-during-coronavirus-covid-19/the-visitor-economy#shops-4-1)  [Visitor economy 2.2](https://www.gov.uk/guidance/working-safely-during-coronavirus-covid-19/the-visitor-economy#shops-4-2)  [Visitor economy 4.2](https://www.gov.uk/guidance/working-safely-during-coronavirus-covid-19/the-visitor-economy#shops-3-2) |
|  | Reducing maximum occupancy for lifts, providing hand sanitiser for the operation of lifts and encouraging use of stairs wherever possible. | How have you reduced lift occupancy, and have you encouraged the use of stairs? Please provide details. | [Performing arts 3.6](https://www.gov.uk/guidance/working-safely-during-coronavirus-covid-19/performing-arts#arts-3-6)  [Visitor economy 2.1](https://www.gov.uk/guidance/working-safely-during-coronavirus-covid-19/the-visitor-economy#shops-4-1)  [Visitor economy 4.2](https://www.gov.uk/guidance/working-safely-during-coronavirus-covid-19/the-visitor-economy#shops-3-2) |
|  | Reducing instances where people might be required to queue. | How have you reduced instances where people might be required to queue? | [Performing arts 3.11](https://www.gov.uk/guidance/working-safely-during-coronavirus-covid-19/performing-arts#arts-3-11)  [Restaurants, pubs, bars and takeaway services 2.1](https://www.gov.uk/guidance/working-safely-during-coronavirus-covid-19/restaurants-offering-takeaway-or-delivery#takeaways-2-1)  [Visitor economy 2.1](https://www.gov.uk/guidance/working-safely-during-coronavirus-covid-19/the-visitor-economy#shops-4-1) |
|  | Management of crowd density points, such as where people stop to watch displays, must be considered. | How have you reduced instances where people might be required to queue? | [Performing arts 3.11](https://www.gov.uk/guidance/working-safely-during-coronavirus-covid-19/performing-arts#arts-3-11)  [Visitor economy 2.1](https://www.gov.uk/guidance/working-safely-during-coronavirus-covid-19/the-visitor-economy#shops-4-1) |
| Equality of access | Making sure that people with disabilities are able to access lifts. | How have you ensured that people with disabilities are able to access lifts? | [Performing arts 3.6](https://www.gov.uk/guidance/working-safely-during-coronavirus-covid-19/performing-arts#arts-3-6) |

#### 2.6 Ticketing and payments

| **Key considerations** | **Potential mitigations** | **Suggested questions** | **Relevant guidance** |
| --- | --- | --- | --- |
| Reducing ticket numbers | Enabling a booking system or other approaches (e.g. timed ticketing) to manage demand of spaces. | Have you implemented a booking system to manage demand for spaces? Please provide details. | [Performing arts 3.1](https://www.gov.uk/guidance/working-safely-during-coronavirus-covid-19/performing-arts#arts-3-1)  [Places of worship 5](https://www.gov.uk/government/publications/covid-19-guidance-for-the-safe-use-of-places-of-worship-during-the-pandemic-from-4-july/covid-19-guidance-for-the-safe-use-of-places-of-worship-during-the-pandemic-from-4-july)  [SG02 3.13](https://sgsa.org.uk/wp-content/uploads/2020/08/SG02-Planning-for-Social-Distancing-at-Sports-Grounds.pdf)  [Visitor economy 2.1](https://www.gov.uk/guidance/working-safely-during-coronavirus-covid-19/the-visitor-economy#shops-4-1)  [Visitor economy 2.2](https://www.gov.uk/guidance/working-safely-during-coronavirus-covid-19/the-visitor-economy#shops-4-2) |
|  | Allowing a sufficient break time between sessions or performances held to prevent waiting in groups. | Have you allowed a break between sessions or performances? | [Performing arts 3.1](https://www.gov.uk/guidance/working-safely-during-coronavirus-covid-19/performing-arts#arts-3-1) |
|  | Operating on a book-in-advance basis for any spaces available to hire, preferably online or over the phone. | Have you implemented a book-in-advance basis for spaces for hire? How is this conducted? | [Performing arts 3.1](https://www.gov.uk/guidance/working-safely-during-coronavirus-covid-19/performing-arts#arts-3-1)  [SG02 3.13](https://sgsa.org.uk/wp-content/uploads/2020/08/SG02-Planning-for-Social-Distancing-at-Sports-Grounds.pdf)  [Visitor economy 2.1](https://www.gov.uk/guidance/working-safely-during-coronavirus-covid-19/the-visitor-economy#shops-4-1) |
|  | Encouraging guests to purchase tickets online and to use e-ticketing. Where this is not the case, encouraging contactless payment. | Have you encouraged guests to purchase tickets online and use e-ticketing? | [Performing arts 3.8](https://www.gov.uk/guidance/working-safely-during-coronavirus-covid-19/performing-arts#arts-3-8)  [SG02 3.13](https://sgsa.org.uk/wp-content/uploads/2020/08/SG02-Planning-for-Social-Distancing-at-Sports-Grounds.pdf)  [Visitor economy 4.3](https://www.gov.uk/guidance/working-safely-during-coronavirus-covid-19/the-visitor-economy#shops-3-3) |
|  | Maintaining social distancing as far as possible when checking tickets. | How are you maintaining social distancing when checking tickets? | [Performing arts 3.8](https://www.gov.uk/guidance/working-safely-during-coronavirus-covid-19/performing-arts#arts-3-8) |
|  | Where there is no ticketing, considering using other communications approaches, coupled with stewarding. | If you have no ticketing, how are you managing demand of spaces? | [Performing arts 3.2](https://www.gov.uk/guidance/working-safely-during-coronavirus-covid-19/performing-arts#arts-2-1) |

#### 2.7 Screening procedures

| **Key considerations** | **Potential mitigations** | **Suggested questions** | **Relevant guidance** |
| --- | --- | --- | --- |
| Identifying individuals with potential or confirmed COVID19 | Attendees should not attend if they have symptoms consistent with COVID19, or have been advised to self-isolate by relevant authorities. | Have you advised attendees with potential COVID19 (or those advised to self-isolate) not to attend? | [Performing arts 3.4](https://www.gov.uk/guidance/working-safely-during-coronavirus-covid-19/performing-arts#arts-3-4)  [Places of worship 5](https://www.gov.uk/government/publications/covid-19-guidance-for-the-safe-use-of-places-of-worship-during-the-pandemic-from-4-july/covid-19-guidance-for-the-safe-use-of-places-of-worship-during-the-pandemic-from-4-july)  [Return of sport](https://www.gov.uk/government/publications/coronavirus-covid-19-guidance-on-phased-return-of-sport-and-recreation/guidance-for-the-public-on-the-phased-return-of-outdoor-sport-and-recreation)  [Visitor economy 2.2](https://www.gov.uk/guidance/working-safely-during-coronavirus-covid-19/the-visitor-economy#shops-4-2) |
|  | Screening of anyone prior to entry into venues, which may include, but not be limited to, a COVID19 symptom questionnaire. | How are you screening people prior to entry into venues? | [Performing arts 4.1](https://www.gov.uk/guidance/working-safely-during-coronavirus-covid-19/performing-arts#arts-4-1) |
|  | Consideration should be given to all those coming onto the site – other than the attendees – being required to sign a register or form confirming they are not suffering from COVID19 symptoms or living in the same household or support bubble as someone who is unwell. | How are you screening people prior to entry into venues? | [Events Industry Forum 3.1](https://www.eventindustrynews.com/wp-content/uploads/2020/07/EIfDCMS-COVID-19-Working-Safely-9-July-2020.pdf) |
|  | Temperature checks are unreliable indicators of illness and should not be used. | How are you screening people prior to entry into venues? | [MHRA](https://www.gov.uk/government/news/dont-rely-on-temperature-screening-products-for-detection-of-coronavirus-covid-19-says-mhra) |

### 3 COVID risk domain: Security and emergencies

#### 3.1 Security

| **Key considerations** | **Potential mitigations** | **Suggested questions** | **Relevant guidance** |
| --- | --- | --- | --- |
| General security measures | Operators should try and organise queuing within existing protected areas. | Have queues been organised in relation to the structure of the surrounding areas? | [Hotels and guest accommodation 2.4](https://www.gov.uk/guidance/working-safely-during-coronavirus-covid-19/hotels-and-other-guest-accommodation#section-2-4)  [Visitor economy 2.3](https://www.gov.uk/guidance/working-safely-during-coronavirus-covid-19/the-visitor-economy#shops-4-3) |
|  | Operators should be careful to avoid giving credible, detailed information that could help a hostile entity identify an attractive target and carry out an attack. | How have you ensured that security has been maintained in your venue? | [Hotels and guest accommodation 2.4](https://www.gov.uk/guidance/working-safely-during-coronavirus-covid-19/hotels-and-other-guest-accommodation#section-2-4)  [Visitor economy 2.3](https://www.gov.uk/guidance/working-safely-during-coronavirus-covid-19/the-visitor-economy#shops-4-3) |
|  | Whilst stewards and security officers may be focussed on managing people and queues for COVID19 safety reasons, they should continue to remain vigilant for and report any suspicious activity as soon as possible. | How have you ensured that security has been maintained in your venue? | [Hotels and guest accommodation 2.4](https://www.gov.uk/guidance/working-safely-during-coronavirus-covid-19/hotels-and-other-guest-accommodation#section-2-4)  [Visitor economy 2.3](https://www.gov.uk/guidance/working-safely-during-coronavirus-covid-19/the-visitor-economy#shops-4-3) |
|  | Restricted access entry points, such as those facilitated by keypad, biometrics and/or pass should remain fully in operation. | Have you ensured that restricted access entry points are still in operation? | [Hotels and guest accommodation 2.4](https://www.gov.uk/guidance/working-safely-during-coronavirus-covid-19/hotels-and-other-guest-accommodation#section-2-4)  [Visitor economy 2.3](https://www.gov.uk/guidance/working-safely-during-coronavirus-covid-19/the-visitor-economy#shops-4-3) |
|  | Reviewing external messaging to visitors and audience to make sure it does not provide information that may present a security risk e.g. location or number of people permitted in a queue. | How are you reviewing external messaging to ensure it does not pose a security risk? | [Performing arts 3.14](https://www.gov.uk/guidance/working-safely-during-coronavirus-covid-19/performing-arts#arts-3-14) |
| Emergency procedures | Reviewing your incident and emergency procedures to ensure they reflect and enable the social distancing principles as far as possible. | How have you reviewed your incident and emergency procedures to ensure they reflect and enable the social distancing principles as far as possible? | [Performing arts 4.6](https://www.gov.uk/guidance/working-safely-during-coronavirus-covid-19/performing-arts#arts-4-6)  [Restaurants, pubs, bars and takeaway services 4.8](https://www.gov.uk/guidance/working-safely-during-coronavirus-covid-19/restaurants-offering-takeaway-or-delivery#takeaways-4-8)  [Return of sport](https://www.gov.uk/government/publications/coronavirus-covid-19-guidance-on-phased-return-of-sport-and-recreation/guidance-for-the-public-on-the-phased-return-of-outdoor-sport-and-recreation)  [Visitor economy 2.1](https://www.gov.uk/guidance/working-safely-during-coronavirus-covid-19/the-visitor-economy#shops-4-1)  [Visitor economy 2.3](https://www.gov.uk/guidance/working-safely-during-coronavirus-covid-19/the-visitor-economy#shops-4-3)  [Visitor economy 4.4](https://www.gov.uk/guidance/working-safely-during-coronavirus-covid-19/the-visitor-economy#shops-3-4) |
|  | Considering the security implications of any changes you intend to make to your operations and practices in response to COVID19, as any revisions may present new or altered security risks which may need mitigations. | Have you considered the security implications of COVID-secure changes? Please provide details. | [Heritage](https://www.gov.uk/guidance/working-safely-during-coronavirus-covid-19/heritage-locations)  [Hotels and guest accommodation 2.4](https://www.gov.uk/guidance/working-safely-during-coronavirus-covid-19/hotels-and-other-guest-accommodation#section-2-4)  [Hotels and guest accommodation 4.6](https://www.gov.uk/guidance/working-safely-during-coronavirus-covid-19/hotels-and-other-guest-accommodation#section-4-6)  [Performing arts 4.6](https://www.gov.uk/guidance/working-safely-during-coronavirus-covid-19/performing-arts#arts-4-6)  [Restaurants, pubs, bars and takeaway services 4.8](https://www.gov.uk/guidance/working-safely-during-coronavirus-covid-19/restaurants-offering-takeaway-or-delivery#takeaways-4-8)  [Visitor economy 2.1](https://www.gov.uk/guidance/working-safely-during-coronavirus-covid-19/the-visitor-economy#shops-4-1)  [Visitor economy 2.3](https://www.gov.uk/guidance/working-safely-during-coronavirus-covid-19/the-visitor-economy#shops-4-3) |
|  | Considering whether you have enough appropriately trained staff to keep people safe. | Do you have enough appropriately trained staff to keep people safe? | [Performing arts 4.6](https://www.gov.uk/guidance/working-safely-during-coronavirus-covid-19/performing-arts#arts-4-6)  [Restaurants, pubs, bars and takeaway services 4.8](https://www.gov.uk/guidance/working-safely-during-coronavirus-covid-19/restaurants-offering-takeaway-or-delivery#takeaways-4-8) |
|  | Consider providing separate stewarding to manage the social distancing and other safety aspects to enable your security staff to focus on their core responsibilities to keep the site safe from threats. | Have you designated staff to manage queues? | [Visitor economy 2.3](https://www.gov.uk/guidance/working-safely-during-coronavirus-covid-19/the-visitor-economy#shops-4-3) |
|  | Ensure there is a good communication system in place to inform people of any incident. Carry out a short exercise or test to check procedures and equipment for this are working correctly. | Have you conducted a short test or exercise to check that incident response procedures are working? | [Visitor economy 2.3](https://www.gov.uk/guidance/working-safely-during-coronavirus-covid-19/the-visitor-economy#shops-4-3) |
| Search and screening processes* | Conduct of physical search and screening of staff, contractors and visitors may need adapting in order to adhere to social distancing measures. | How have you adapted physical search and screening of staff, contractors and visitors to adhere to social distancing measures? | [Hotels and guest accommodation 2.4](https://www.gov.uk/guidance/working-safely-during-coronavirus-covid-19/hotels-and-other-guest-accommodation#section-2-4)  [Visitor economy 2.3](https://www.gov.uk/guidance/working-safely-during-coronavirus-covid-19/the-visitor-economy#shops-4-3) |

*The Centre for the Protection of National Infrastructure (CPNI) has published guidance on adapting existing search and screening processes to take account of physical distancing.

#### 3.2 In case of suspected COVID cases

| **Key considerations** | **Potential mitigations** | **Suggested questions** | **Relevant guidance** |
| --- | --- | --- | --- |
| Immediate management of suspected COVID cases | Ensuring there is a clear policy in place for managing a COVID19 suspected or confirmed individual, and abiding by government and PHE guidelines and reporting requirements. | What policy do you have in place for managing a COVID19 suspected or confirmed individual? Please provide details. | [Performing arts 4.1](https://www.gov.uk/guidance/working-safely-during-coronavirus-covid-19/performing-arts#arts-4-1) |
|  | Consideration should be given to creating an isolation/quarantine point, close to the entrance or exit, where anyone found to be unwell or at risk can be taken. | Have you created an isolation/quarantine point where anyone found to be unwell or at risk can be taken? Please provide details. | [Events Industry Forum 3.1](https://www.eventindustrynews.com/wp-content/uploads/2020/07/EIfDCMS-COVID-19-Working-Safely-9-July-2020.pdf) |
|  | Consideration should be given to creating an isolation/quarantine area (ideally close to medical facilities) from the start of construction through to the conclusion of breakdown. | Have you created an isolation/quarantine point where anyone found to be unwell or at risk can be taken? Please provide details. | [Events Industry Forum 3.2](https://www.eventindustrynews.com/wp-content/uploads/2020/07/EIfDCMS-COVID-19-Working-Safely-9-July-2020.pdf) |
|  | If you need to provide assistance to an individual who is symptomatic and may have COVID19, wherever possible, place the person in a place away from others. If there is no physically separate room, ask others who are not involved in providing assistance to stay at least 2 metres away from the individual. If barriers or screens are available, these may be used. | Is there a policy in place for managing a COVID19 suspected or confirmed individual? Please provide details. | [Guidance for first responders](https://www.gov.uk/government/publications/novel-coronavirus-2019-ncov-interim-guidance-for-first-responders/interim-guidance-for-first-responders-and-others-in-close-contact-with-symptomatic-people-with-potential-2019-ncov) |
| Management of suspected COVID in accommodation | Accommodation providers should consider how they would manage a situation with an unwell guest, including whether symptomatic guests in self-isolation would be responsible for cleaning their own rooms and stripping/making their own beds. | Is there a policy in place for managing a COVID19 suspected or confirmed individual? Please provide details. | [Hotels and guest accommodation 5.2](https://www.gov.uk/guidance/working-safely-during-coronavirus-covid-19/hotels-and-other-guest-accommodation#section-5-2) |
|  | Where an accommodation provider has a COVID-symptomatic guest, they should agree next steps with the guest at the earliest opportunity, ensuring no onward risk of infection to other guests or workers. | Is there a policy in place for managing a COVID19 suspected or confirmed individual? Please provide details. | [Hotels and guest accommodation 5.2](https://www.gov.uk/guidance/working-safely-during-coronavirus-covid-19/hotels-and-other-guest-accommodation#section-5-2) |

#### 3.3 Compliance with contact tracing requirements

| **Key considerations** | **Potential mitigations** | **Suggested questions** | **Relevant guidance** |
| --- | --- | --- | --- |
| Following national guidance around ensuring attendees and staff can be contact traced if required* | Keeping a temporary record of your attendees (when applicable) and other visitors for 21 days. | How are you storing details of your attendees in order to remain compliant with contact tracing requirements? | [Heritage](https://www.gov.uk/guidance/working-safely-during-coronavirus-covid-19/heritage-locations)  [Performing arts 1.3](https://www.gov.uk/guidance/working-safely-during-coronavirus-covid-19/performing-arts#arts-1-3)  [Places of worship 5](https://www.gov.uk/government/publications/covid-19-guidance-for-the-safe-use-of-places-of-worship-during-the-pandemic-from-4-july/covid-19-guidance-for-the-safe-use-of-places-of-worship-during-the-pandemic-from-4-july)  [Maintaining records](https://www.gov.uk/guidance/maintaining-records-of-staff-customers-and-visitors-to-support-nhs-test-and-trace) |
|  | Collect contact details for all audience members, in addition to the lead booker, and recording where audience members are seated. | How are you storing details of your attendees in order to remain compliant with contact tracing requirements? | [Heritage](https://www.gov.uk/guidance/working-safely-during-coronavirus-covid-19/heritage-locations)  [Performing arts 1.3](https://www.gov.uk/guidance/working-safely-during-coronavirus-covid-19/performing-arts#arts-1-3)  [Places of worship 5](https://www.gov.uk/government/publications/covid-19-guidance-for-the-safe-use-of-places-of-worship-during-the-pandemic-from-4-july/covid-19-guidance-for-the-safe-use-of-places-of-worship-during-the-pandemic-from-4-july)  [Maintaining records](https://www.gov.uk/guidance/maintaining-records-of-staff-customers-and-visitors-to-support-nhs-test-and-trace) |
|  | Physios or their equivalent, should keep a record of each participant they have come into contact with for test and trace purposes. | How are you storing details of your attendees in order to remain compliant with contact tracing requirements? | [Return of sport](https://www.gov.uk/government/publications/coronavirus-covid-19-guidance-on-phased-return-of-sport-and-recreation/guidance-for-the-public-on-the-phased-return-of-outdoor-sport-and-recreation)  [Maintaining records](https://www.gov.uk/guidance/maintaining-records-of-staff-customers-and-visitors-to-support-nhs-test-and-trace) |
|  | Keeping a temporary record of staff shift patterns for 21 days and assist NHS Test and Trace with requests for that data if needed. | How are you storing details of your staff in order to remain compliant with contact tracing requirements? | [Heritage](https://www.gov.uk/guidance/working-safely-during-coronavirus-covid-19/heritage-locations)  [Visitor economy 7.1](https://www.gov.uk/guidance/working-safely-during-coronavirus-covid-19/the-visitor-economy#shops-7-1) |
|  | You must register for an official NHS QR code and display the official NHS QR poster. | Have you registered for an official NHS QR code and displayed the official NHS QR poster? | [Heritage](https://www.gov.uk/guidance/working-safely-during-coronavirus-covid-19/heritage-locations)  [Maintaining records](https://www.gov.uk/guidance/maintaining-records-of-staff-customers-and-visitors-to-support-nhs-test-and-trace)  [Places of worship 5](https://www.gov.uk/government/publications/covid-19-guidance-for-the-safe-use-of-places-of-worship-during-the-pandemic-from-4-july/covid-19-guidance-for-the-safe-use-of-places-of-worship-during-the-pandemic-from-4-july) |

*Further details available in the [maintaining records](https://www.gov.uk/guidance/maintaining-records-of-staff-customers-and-visitors-to-support-nhs-test-and-trace) guidance.

### 4 COVID risk domain: Information for guests and workers

| **Key considerations** | **Potential mitigations** | **Suggested questions** | **Relevant guidance** |
| --- | --- | --- | --- |
| Self-isolation | Communicating ahead of arrival and on arrival the guidance about who should self-isolate. | How are you communicating the guidance about who should self-isolate ahead of arrival? | [Performing arts 3.5](https://www.gov.uk/guidance/working-safely-during-coronavirus-covid-19/performing-arts#arts-3-5) |
| Signage and communication | Providing written or spoken communication of the latest guidelines to both workers and customers inside and outside the venue. | How are you providing communication of the latest guidelines to workers and customers inside and outside the venue? | [Food businesses](https://www.gov.uk/government/publications/covid-19-guidance-for-food-businesses/guidance-for-food-businesses-on-coronavirus-covid-19)  [Hotels and guest accommodation 2.3](https://www.gov.uk/guidance/working-safely-during-coronavirus-covid-19/hotels-and-other-guest-accommodation#section-2-3)  [Performing arts 3.14](https://www.gov.uk/guidance/working-safely-during-coronavirus-covid-19/performing-arts#arts-3-14)  [Restaurants, pubs, bars and takeaway services 2.5](https://www.gov.uk/guidance/working-safely-during-coronavirus-covid-19/restaurants-offering-takeaway-or-delivery#takeaways-2-5)  [Return of sport](https://www.gov.uk/government/publications/coronavirus-covid-19-guidance-on-phased-return-of-sport-and-recreation/guidance-for-the-public-on-the-phased-return-of-outdoor-sport-and-recreation)  [SG02 3.12](https://sgsa.org.uk/wp-content/uploads/2020/08/SG02-Planning-for-Social-Distancing-at-Sports-Grounds.pdf)  [Visitor economy 2.1](https://www.gov.uk/guidance/working-safely-during-coronavirus-covid-19/the-visitor-economy#shops-4-1)  [Visitor economy 2.2](https://www.gov.uk/guidance/working-safely-during-coronavirus-covid-19/the-visitor-economy#shops-4-2) |
|  | Using signage (for example, posters or leaflets on basic hygiene practices such as handwashing) in each room. | What COVID19 signage are you providing in each room? | [Food businesses](https://www.gov.uk/government/publications/covid-19-guidance-for-food-businesses/guidance-for-food-businesses-on-coronavirus-covid-19)  [Hotels and guest accommodation 2.3](https://www.gov.uk/guidance/working-safely-during-coronavirus-covid-19/hotels-and-other-guest-accommodation#section-2-3)  [Places of worship 4](https://www.gov.uk/government/publications/covid-19-guidance-for-the-safe-use-of-places-of-worship-during-the-pandemic-from-4-july/covid-19-guidance-for-the-safe-use-of-places-of-worship-during-the-pandemic-from-4-july)  [SG02 3.12](https://sgsa.org.uk/wp-content/uploads/2020/08/SG02-Planning-for-Social-Distancing-at-Sports-Grounds.pdf)  [Visitor economy 2.1](https://www.gov.uk/guidance/working-safely-during-coronavirus-covid-19/the-visitor-economy#shops-4-1)  [Visitor economy 2.2](https://www.gov.uk/guidance/working-safely-during-coronavirus-covid-19/the-visitor-economy#shops-4-2) |
|  | Providing clear guidance both before arrival and on arrival, for example through signage, visual aids, on your website and in pre-arrival emails, and on email confirmation when purchasing tickets. | How are you communicating guidance to guests before arrival and on arrival? | [Hotels and guest accommodation 2.3](https://www.gov.uk/guidance/working-safely-during-coronavirus-covid-19/hotels-and-other-guest-accommodation#section-2-3)  [Performing arts 3.14](https://www.gov.uk/guidance/working-safely-during-coronavirus-covid-19/performing-arts#arts-3-14)  [Places of worship 4](https://www.gov.uk/government/publications/covid-19-guidance-for-the-safe-use-of-places-of-worship-during-the-pandemic-from-4-july/covid-19-guidance-for-the-safe-use-of-places-of-worship-during-the-pandemic-from-4-july)  [Restaurants, pubs, bars and takeaway services 2.5](https://www.gov.uk/guidance/working-safely-during-coronavirus-covid-19/restaurants-offering-takeaway-or-delivery#takeaways-2-5)  [Return of sport](https://www.gov.uk/government/publications/coronavirus-covid-19-guidance-on-phased-return-of-sport-and-recreation/guidance-for-the-public-on-the-phased-return-of-outdoor-sport-and-recreation)  [Visitor economy 2.1](https://www.gov.uk/guidance/working-safely-during-coronavirus-covid-19/the-visitor-economy#shops-4-1)  [Visitor economy 2.2](https://www.gov.uk/guidance/working-safely-during-coronavirus-covid-19/the-visitor-economy#shops-4-2) |
|  | Informing guests of government guidance on face coverings. | How are you informing guests of government guidance on face coverings? | [Hotels and guest accommodation 2.3](https://www.gov.uk/guidance/working-safely-during-coronavirus-covid-19/hotels-and-other-guest-accommodation#section-2-3)  [Places of worship 5](https://www.gov.uk/government/publications/covid-19-guidance-for-the-safe-use-of-places-of-worship-during-the-pandemic-from-4-july/covid-19-guidance-for-the-safe-use-of-places-of-worship-during-the-pandemic-from-4-july)  [Visitor economy 2.1](https://www.gov.uk/guidance/working-safely-during-coronavirus-covid-19/the-visitor-economy#shops-4-1) |
|  | Reminding guests of social distancing guidelines during check-in. | How are you reminding guests of social distancing guidelines during check-in? | [Hotels and guest accommodation 2.3](https://www.gov.uk/guidance/working-safely-during-coronavirus-covid-19/hotels-and-other-guest-accommodation#section-2-3) |
|  | Attendees who are accompanied by children should be reminded that they are responsible for supervising them at all times and should follow social distancing guidelines. | How are you reminding guests of social distancing guidelines during check-in? | [Events Industry Forum 4.3](https://www.eventindustrynews.com/wp-content/uploads/2020/07/EIfDCMS-COVID-19-Working-Safely-9-July-2020.pdf)  [Places of worship 5](https://www.gov.uk/government/publications/covid-19-guidance-for-the-safe-use-of-places-of-worship-during-the-pandemic-from-4-july/covid-19-guidance-for-the-safe-use-of-places-of-worship-during-the-pandemic-from-4-july) |
|  | Where necessary, informing customers that police and the local authorities have the powers to enforce requirements in relation to social distancing and may instruct customers to disperse, leave an area, issue a fixed penalty notice or take further enforcement action. | How are you reminding guests of social distancing guidelines during check-in? | [Places of worship 5](https://www.gov.uk/government/publications/covid-19-guidance-for-the-safe-use-of-places-of-worship-during-the-pandemic-from-4-july/covid-19-guidance-for-the-safe-use-of-places-of-worship-during-the-pandemic-from-4-july)  [Restaurants, pubs, bars and takeaway services 2.5](https://www.gov.uk/guidance/working-safely-during-coronavirus-covid-19/restaurants-offering-takeaway-or-delivery#takeaways-2-5) |
|  | Frequent updates of communication (weekly is suggested) to prevent it becoming stale. Regular communication, even if there is little new to share, is important to reinforce key messages and prevent false information from circulating. | How frequently are you providing updates of communication to guests and workers? | [Food businesses](https://www.gov.uk/government/publications/covid-19-guidance-for-food-businesses/guidance-for-food-businesses-on-coronavirus-covid-19)  [Visitor economy 2.1](https://www.gov.uk/guidance/working-safely-during-coronavirus-covid-19/the-visitor-economy#shops-4-1)  [Visitor economy 7.3](https://www.gov.uk/guidance/working-safely-during-coronavirus-covid-19/the-visitor-economy#shops-7-3) |
| Equality of access | Considering the equalities impacts of the changes made and what advice or guidance you will need to provide for users who might be adversely impacted. | How have you provided guidance or advice for users with disabilities who may be adversely impacted? | [Performing arts 3.14](https://www.gov.uk/guidance/working-safely-during-coronavirus-covid-19/performing-arts#arts-3-14)  [Places of worship](https://www.gov.uk/government/publications/covid-19-guidance-for-the-safe-use-of-places-of-worship-during-the-pandemic-from-4-july/covid-19-guidance-for-the-safe-use-of-places-of-worship-during-the-pandemic-from-4-july)  [Visitor economy 2.1](https://www.gov.uk/guidance/working-safely-during-coronavirus-covid-19/the-visitor-economy#shops-4-1)  [Visitor economy 7.3](https://www.gov.uk/guidance/working-safely-during-coronavirus-covid-19/the-visitor-economy#shops-7-3) |
|  | Consider the particular needs of those with protected characteristics, such as people with visual impairments, when developing communication. | How have you accommodated the needs of those with protected characteristics when developing communication? | [Hotels and guest accommodation 2.3](https://www.gov.uk/guidance/working-safely-during-coronavirus-covid-19/hotels-and-other-guest-accommodation#section-2-3)  [Visitor economy 2.1](https://www.gov.uk/guidance/working-safely-during-coronavirus-covid-19/the-visitor-economy#shops-4-1)  [Visitor economy 7.3](https://www.gov.uk/guidance/working-safely-during-coronavirus-covid-19/the-visitor-economy#shops-7-3) |
|  | Consider using easy-to-translate communication materials which can be translated, using free-to-use online translation services, into the preferred languages of employees if English is not their first language. | How have you accommodated the needs of those with protected characteristics when developing communication? | [Food businesses](https://www.gov.uk/government/publications/covid-19-guidance-for-food-businesses/guidance-for-food-businesses-on-coronavirus-covid-19)  [Visitor economy 7.3](https://www.gov.uk/guidance/working-safely-during-coronavirus-covid-19/the-visitor-economy#shops-7-3) |

### 5 COVID risk domain: Staff and workforce

Please note that this guidance does not include the mitigations or other modifications required as part of the rehearsal or planning process, only those involved in the running of an event or function on the day itself.

Furthermore, while worker-specific mitigations are listed in this section, those listed elsewhere (including in “sanitation facilities” and “crowd control”) should also be considered as applying to staff.

#### 5.1  Movement of staff

| **Key considerations** | **Potential mitigations** | **Suggested questions** | **Relevant guidance** |
| --- | --- | --- | --- |
| Minimising movement | Reducing movement by discouraging non-essential trips within buildings and sites. | How have you reduced non-essential trips for staff within buildings and sites? | [Performing arts 3.6](https://www.gov.uk/guidance/working-safely-during-coronavirus-covid-19/performing-arts#arts-3-6)  [Restaurants, pubs, bars and takeaway services 4.1](https://www.gov.uk/guidance/working-safely-during-coronavirus-covid-19/restaurants-offering-takeaway-or-delivery#takeaways-4-1)  [Visitor economy 4.2](https://www.gov.uk/guidance/working-safely-during-coronavirus-covid-19/the-visitor-economy#shops-3-2) |
|  | Consider the maximum number of people who can be safely accommodated on site. | What is the maximum number of people who can safely be accommodated on site? | [Hotels and guest accommodation 3](https://www.gov.uk/guidance/working-safely-during-coronavirus-covid-19/hotels-and-other-guest-accommodation#shops-3-1)  [Visitor economy 3](https://www.gov.uk/guidance/working-safely-during-coronavirus-covid-19/the-visitor-economy#shops-5) |
|  | Limit unnecessary visits to the site. | How have you reduced non-essential trips for staff within buildings and sites? | [Food businesses](https://www.gov.uk/government/publications/covid-19-guidance-for-food-businesses/guidance-for-food-businesses-on-coronavirus-covid-19) |
|  | Reducing congestion, for example by having more entry points to the workplace. If you have more than one door, consider having one for entering the building and one for exiting. | How have you reduced congestion in the workplace? | [Restaurants, pubs, bars and takeaway services 4.1](https://www.gov.uk/guidance/working-safely-during-coronavirus-covid-19/restaurants-offering-takeaway-or-delivery#takeaways-4-1)  [Visitor economy 4.1](https://www.gov.uk/guidance/working-safely-during-coronavirus-covid-19/the-visitor-economy#shops-3-1) |
|  | Using markings to guide staff coming into or leaving the building. | How have you used markings to guide staff entering or leaving the building? | [Restaurants, pubs, bars and takeaway services 4.1](https://www.gov.uk/guidance/working-safely-during-coronavirus-covid-19/restaurants-offering-takeaway-or-delivery#takeaways-4-1)  [Visitor economy 4.1](https://www.gov.uk/guidance/working-safely-during-coronavirus-covid-19/the-visitor-economy#shops-3-1) |

#### 5.2 Exposure

| **Key considerations** | **Potential mitigations** | **Suggested questions** | **Relevant guidance** |
| --- | --- | --- | --- |
| Minimising interaction with attendees | Ensuring that members of fixed teams are particularly careful to maintain social distancing when interacting with audience members. | How are you maintaining social distancing between staff and audience members? | [Performing arts 3.7](https://www.gov.uk/guidance/working-safely-during-coronavirus-covid-19/performing-arts#arts-3-7) |
|  | Identifying any roles that interact with audiences and manage transmission risk appropriately. | How are you reducing transmission risk between staff and attendees? | [Performing arts 3.7](https://www.gov.uk/guidance/working-safely-during-coronavirus-covid-19/performing-arts#arts-3-7) |
|  | Limiting the potential for guest contact with performers and support staff. | How are you reducing transmission risk between staff and attendees? | [Performing arts 3.11](https://www.gov.uk/guidance/working-safely-during-coronavirus-covid-19/performing-arts#arts-3-11) |
|  | Considering using booths, barriers or screens between performers and any audience, with comprehensive risk assessments to ensure that transmission risk is appropriately contained and that other health and safety risks are managed. | Please outline what mitigations you are using to minimise transmission risk between staff and attendees. | [Performing arts 4.1](https://www.gov.uk/guidance/working-safely-during-coronavirus-covid-19/performing-arts#arts-4-1)  [Visitor economy 4.3](https://www.gov.uk/guidance/working-safely-during-coronavirus-covid-19/the-visitor-economy#shops-3-3) |
| Minimising interaction with other employees | Consider creating cohorts or groups of staff to minimise contact and reduce potential transmission. Cleaning should also be scheduled around zones and cohort lines. | Have you created cohorts or groups of staff? | [Food businesses](https://www.gov.uk/government/publications/covid-19-guidance-for-food-businesses/guidance-for-food-businesses-on-coronavirus-covid-19)  [Performing arts 4.8](https://www.gov.uk/guidance/working-safely-during-coronavirus-covid-19/performing-arts#arts-4-8)  [Restaurants, pubs, bars and takeaway services 4.1](https://www.gov.uk/guidance/working-safely-during-coronavirus-covid-19/restaurants-offering-takeaway-or-delivery#takeaways-4-1)  [Visitor economy 7.1](https://www.gov.uk/guidance/working-safely-during-coronavirus-covid-19/the-visitor-economy#shops-7-1) |
|  | Consider staggering shift starting times to minimise crowding at entry points. | Have you staggered shift starting times? | [Food businesses](https://www.gov.uk/government/publications/covid-19-guidance-for-food-businesses/guidance-for-food-businesses-on-coronavirus-covid-19)  [Restaurants, pubs, bars and takeaway services 4.1](https://www.gov.uk/guidance/working-safely-during-coronavirus-covid-19/restaurants-offering-takeaway-or-delivery#takeaways-4-1)  [Places of worship 5](https://www.gov.uk/government/publications/covid-19-guidance-for-the-safe-use-of-places-of-worship-during-the-pandemic-from-4-july/covid-19-guidance-for-the-safe-use-of-places-of-worship-during-the-pandemic-from-4-july)  [Visitor economy 4.1](https://www.gov.uk/guidance/working-safely-during-coronavirus-covid-19/the-visitor-economy#shops-3-1) |
|  | Consider designating managers or senior staff to act as visible marshals to supervise entry points. | Have you designated marshals to supervise entry points for staff? | [Food businesses](https://www.gov.uk/government/publications/covid-19-guidance-for-food-businesses/guidance-for-food-businesses-on-coronavirus-covid-19) |
|  | Consider creating a break between shifts supported by visible marshalling, to minimise overlap and to enable effective cleaning of the working areas | Have you created a break between shifts? Is this supported by marshalling? | [Food businesses](https://www.gov.uk/government/publications/covid-19-guidance-for-food-businesses/guidance-for-food-businesses-on-coronavirus-covid-19) |
|  | Ensure social distancing of 2 metres if possible while awaiting entry, and if not ensure maximum feasible social distancing is practised. | How are you ensuring social distancing is maintained while staff are awaiting entry? | [Food businesses](https://www.gov.uk/government/publications/covid-19-guidance-for-food-businesses/guidance-for-food-businesses-on-coronavirus-covid-19) |
|  | Reviewing layouts and processes to allow staff to work further apart from each other. | How have you modified layouts and processes to allow staff to work further apart? | [Places of worship 5](https://www.gov.uk/government/publications/covid-19-guidance-for-the-safe-use-of-places-of-worship-during-the-pandemic-from-4-july/covid-19-guidance-for-the-safe-use-of-places-of-worship-during-the-pandemic-from-4-july)  [Restaurants, pubs, bars and takeaway services 4.3](https://www.gov.uk/guidance/working-safely-during-coronavirus-covid-19/restaurants-offering-takeaway-or-delivery#takeaways-4-3)  [Visitor economy 4.3](https://www.gov.uk/guidance/working-safely-during-coronavirus-covid-19/the-visitor-economy#shops-3-3) |
|  | Minimising contact at ‘handover’ points with other staff, such as when presenting food to serving staff and delivery drivers. | How have you minimised contact at ‘handover’ points? | [Restaurants, pubs, bars and takeaway services 4.4](https://www.gov.uk/guidance/working-safely-during-coronavirus-covid-19/restaurants-offering-takeaway-or-delivery#takeaways-4-4) |
| Hygiene precautions | Hand hygiene should be promoted at break times and between shifts. | How are you promoting hand hygiene during breaks and shift changes? | [Restaurants, pubs, bars and takeaway services 4.1](https://www.gov.uk/guidance/working-safely-during-coronavirus-covid-19/restaurants-offering-takeaway-or-delivery#takeaways-4-1)  [Food businesses](https://www.gov.uk/government/publications/covid-19-guidance-for-food-businesses/guidance-for-food-businesses-on-coronavirus-covid-19)  [Visitor economy 4.1](https://www.gov.uk/guidance/working-safely-during-coronavirus-covid-19/the-visitor-economy#shops-3-1) |

#### 5.3 Suspected or confirmed cases in staff

| **Key considerations** | **Potential mitigations** | **Suggested questions** | **Relevant guidance** |
| --- | --- | --- | --- |
| Staff with suspected or confirmed COVID-19 | Employers must not knowingly require or encourage someone who is being required to self-isolate to come to work. | Have you ensured that staff with suspected or confirmed COVID-19 are not required to come to work? | [Restaurants, pubs, bars and takeaway services](https://www.gov.uk/guidance/working-safely-during-coronavirus-covid-19/restaurants-offering-takeaway-or-delivery) |

#### 5.4 Equality of access

| **Key considerations** | **Potential mitigations** | **Suggested questions** | **Relevant guidance** |
| --- | --- | --- | --- |
| Ensuring measures do not negatively impact workers with protected characteristics | Making reasonable adjustments to avoid disabled workers being put at a disadvantage, and assessing the health and safety risks for new or expectant mothers. | How are you making adjustments to ensure that disabled workers are not disadvantaged? | [Hotels and guest accommodation 3.3](https://www.gov.uk/guidance/working-safely-during-coronavirus-covid-19/hotels-and-other-guest-accommodation#shops-3-3)  [Places of worship 5](https://www.gov.uk/government/publications/covid-19-guidance-for-the-safe-use-of-places-of-worship-during-the-pandemic-from-4-july/covid-19-guidance-for-the-safe-use-of-places-of-worship-during-the-pandemic-from-4-july)  [Visitor economy 3.2](https://www.gov.uk/guidance/working-safely-during-coronavirus-covid-19/the-visitor-economy#shops-5-2) |
|  | Making sure that people with disabilities are able to access lifts. | How are you ensuring that people with disabilities are able to access lifts? | [Restaurants, pubs, bars and takeaway services 4.2](https://www.gov.uk/guidance/working-safely-during-coronavirus-covid-19/restaurants-offering-takeaway-or-delivery#takeaways-4-2) |

#### 5.5 Personal protective equipment (PPE)

| **Key considerations** | **Potential mitigations** | **Suggested questions** | **Relevant guidance** |
| --- | --- | --- | --- |
| PPE during general work | Ensure staff are dressed in an agreed and approved manner. Selection of appropriate PPE and work wear is the employer’s responsibility and representatives and workers should be consulted on selection. | Please detail personal protective equipment (PPE) worn by staff during general work. | [Food businesses](https://www.gov.uk/government/publications/covid-19-guidance-for-food-businesses/guidance-for-food-businesses-on-coronavirus-covid-19) |
| PPE during general cleaning | When cleaning surfaces, it is not necessary to wear PPE or clothing over and above what would usually be used. | Please detail personal protective equipment (PPE) worn by staff during general cleaning. | [Cleaning (non-healthcare settings)](https://www.gov.uk/government/publications/covid-19-decontamination-in-non-healthcare-settings/covid-19-decontamination-in-non-healthcare-settings) |
| PPE during suspected or confirmed COVID19 infection | The minimum PPE to be worn for cleaning an area after a person with symptoms of, or confirmed COVID19 has left the setting possible is disposable gloves and an apron. Wash hands with soap and water for 20 seconds after all PPE has been removed. | Please detail personal protective equipment (PPE) worn by staff during cleaning during suspected or confirmed COVID19 infection. | [Cleaning (non-healthcare settings)](https://www.gov.uk/government/publications/covid-19-decontamination-in-non-healthcare-settings/covid-19-decontamination-in-non-healthcare-settings) |
|  | If a risk assessment of the setting indicates that a higher level of virus may be present (for example, where someone unwell has spent the night such as in a hotel room or boarding school dormitory) then additional PPE to protect the cleaner’s eyes, mouth and nose may be necessary. The local Public Health England (PHE) Health Protection Team can advise on this. | Please detail personal protective equipment (PPE) worn by staff during cleaning during suspected or confirmed COVID19 infection. | [Cleaning (non-healthcare settings)](https://www.gov.uk/government/publications/covid-19-decontamination-in-non-healthcare-settings/covid-19-decontamination-in-non-healthcare-settings) |

#### 5.6 Employer-provided transport

Only applies where the employer is providing transport for staff during the event.

| **Key considerations** | **Potential mitigations** | **Suggested questions** | **Relevant guidance** |
| --- | --- | --- | --- |
| Occupancy | Minimise the number of workers in each vehicle. Consider multiple trips with fewer persons. | How have you reduced the number of occupants per vehicle? | [Food businesses](https://www.gov.uk/government/publications/covid-19-guidance-for-food-businesses/guidance-for-food-businesses-on-coronavirus-covid-19)  [Visitor economy 4.1](https://www.gov.uk/guidance/working-safely-during-coronavirus-covid-19/the-visitor-economy#shops-3-1) |
|  | Consider cohorting workers who share accommodation and work area and transporting them in their cohort. | Are workers being transported in their work cohorts? | [Food businesses](https://www.gov.uk/government/publications/covid-19-guidance-for-food-businesses/guidance-for-food-businesses-on-coronavirus-covid-19) |
| Social distancing | Recommend that face coverings are worn in shared transport as required for public transport. | Have you recommended that face coverings are worn in shared transport? | [Food businesses](https://www.gov.uk/government/publications/covid-19-guidance-for-food-businesses/guidance-for-food-businesses-on-coronavirus-covid-19) |
| Screening for symptoms of COVID19 | Inform workers that if unwell they should stay at home and should not use shared transport or go to work. | Have you informed workers that they should not go to work or used shared transport if unwell? | [Food businesses](https://www.gov.uk/government/publications/covid-19-guidance-for-food-businesses/guidance-for-food-businesses-on-coronavirus-covid-19) |
|  | Consider using visible marshals to ensure unwell staff do not board shared transport. | Are you using marshals to ensure unwell staff do not board transport? | [Food businesses](https://www.gov.uk/government/publications/covid-19-guidance-for-food-businesses/guidance-for-food-businesses-on-coronavirus-covid-19) |
|  | Anyone showing symptoms of COVID19 in transit should be taken off the shared transport, returned to their accommodation and be supported to follow the stay at home guidance. | How are you planning to manage anyone showing symptoms of COVID19 in transit? | [Food businesses](https://www.gov.uk/government/publications/covid-19-guidance-for-food-businesses/guidance-for-food-businesses-on-coronavirus-covid-19) |
| Cleaning | Cleaning shared vehicles between shifts or on handover. | How are you planning to clean shared vehicles? | [Visitor economy 7.2](https://www.gov.uk/guidance/working-safely-during-coronavirus-covid-19/the-visitor-economy#shops-7-2) |

### 6 COVID risk domain: Sanitation and cleaning

#### 6.1 General cleaning procedures

| **Key considerations** | **Potential mitigations** | **Suggested questions** | **Relevant guidance** |
| --- | --- | --- | --- |
| Cleaning procedures upon reopening | Establishing new cleaning regimes for the premises or venue and determining how they will be delivered effectively with the planned hours of operation, for example on a daily basis, with some surfaces cleaned regularly throughout the day. | Please outline new cleaning regimes and frequency for the premise, and how they will be delivered during planned hours of operation. | [Hotels and guest accommodation 5.1](https://www.gov.uk/guidance/working-safely-during-coronavirus-covid-19/hotels-and-other-guest-accommodation#section-5-1)  [Performing arts 5.1](https://www.gov.uk/guidance/working-safely-during-coronavirus-covid-19/performing-arts#arts-5-1)  [Places of worship 4](https://www.gov.uk/government/publications/covid-19-guidance-for-the-safe-use-of-places-of-worship-during-the-pandemic-from-4-july/covid-19-guidance-for-the-safe-use-of-places-of-worship-during-the-pandemic-from-4-july) |
|  | Checking whether you need to service or adjust ventilation systems, for example, so that they do not automatically reduce ventilation levels due to lower than normal occupancy levels. | Have you serviced or adjusted ventilation systems if these are available? | [Hotels and guest accommodation 5.1](https://www.gov.uk/guidance/working-safely-during-coronavirus-covid-19/hotels-and-other-guest-accommodation#section-5-1)  [Performing arts 5.1](https://www.gov.uk/guidance/working-safely-during-coronavirus-covid-19/performing-arts#arts-5-1)  [Restaurants, pubs, bars and takeaway services 5.1](https://www.gov.uk/guidance/working-safely-during-coronavirus-covid-19/restaurants-offering-takeaway-or-delivery#takeaways-5-1)  [Visitor economy 5.1](https://www.gov.uk/guidance/working-safely-during-coronavirus-covid-19/the-visitor-economy#shops-5-1) |
|  | Using any natural ventilation systems such as doors (except fire doors) and windows where feasible to ventilate enclosed space. | How have you ensured ventilation of enclosed spaces? | [Hotels and guest accommodation 5.1](https://www.gov.uk/guidance/working-safely-during-coronavirus-covid-19/hotels-and-other-guest-accommodation#section-5-1)  [Performing arts 5.1](https://www.gov.uk/guidance/working-safely-during-coronavirus-covid-19/performing-arts#arts-5-1)  [Visitor economy 5.2](https://www.gov.uk/guidance/working-safely-during-coronavirus-covid-19/the-visitor-economy#shops-5-2) |
| Keeping the environment clean | Frequent cleaning during events, particularly of touchpoints like door handles and areas which are likely to be used extensively, such as toilets. | How often are you cleaning general premises during events? | [Hotels and guest accommodation 5.2](https://www.gov.uk/guidance/working-safely-during-coronavirus-covid-19/hotels-and-other-guest-accommodation#section-5-2)  [Places of worship 4](https://www.gov.uk/government/publications/covid-19-guidance-for-the-safe-use-of-places-of-worship-during-the-pandemic-from-4-july/covid-19-guidance-for-the-safe-use-of-places-of-worship-during-the-pandemic-from-4-july)  [Performing arts 5.2](https://www.gov.uk/guidance/working-safely-during-coronavirus-covid-19/performing-arts#arts-5-2)  [Visitor economy 5.2](https://www.gov.uk/guidance/working-safely-during-coronavirus-covid-19/the-visitor-economy#shops-5-2) |
|  | Frequent cleaning of work areas and equipment between use, using your usual cleaning products. | How often are you cleaning work areas and equipment between uses? | [Hotels and guest accommodation 5.2](https://www.gov.uk/guidance/working-safely-during-coronavirus-covid-19/hotels-and-other-guest-accommodation#section-5-2)  [Performing arts 5.2](https://www.gov.uk/guidance/working-safely-during-coronavirus-covid-19/performing-arts#arts-5-2)  [Visitor economy 5.2](https://www.gov.uk/guidance/working-safely-during-coronavirus-covid-19/the-visitor-economy#shops-5-2) |
|  | Frequent cleaning of objects and surfaces that are touched regularly. | How often are you cleaning objects and surfaces touched regularly? | [Hotels and guest accommodation 5.2](https://www.gov.uk/guidance/working-safely-during-coronavirus-covid-19/hotels-and-other-guest-accommodation#section-5-2)  [Performing arts 5.2](https://www.gov.uk/guidance/working-safely-during-coronavirus-covid-19/performing-arts#arts-5-2)  [Places of worship 5](https://www.gov.uk/government/publications/covid-19-guidance-for-the-safe-use-of-places-of-worship-during-the-pandemic-from-4-july/covid-19-guidance-for-the-safe-use-of-places-of-worship-during-the-pandemic-from-4-july)  [Restaurants, pubs, bars and takeaway services 5.2](https://www.gov.uk/guidance/working-safely-during-coronavirus-covid-19/restaurants-offering-takeaway-or-delivery#takeaways-5-2) |
|  | Extra, frequent deep cleaning of shared spaces such as audition spaces, rehearsal and backstage areas. | How often are you cleaning shared spaces (e.g. audition spaces, rehearsal areas)? | [Performing arts 5.2](https://www.gov.uk/guidance/working-safely-during-coronavirus-covid-19/performing-arts#arts-5-2) |
| Laundry | Items should be washed in accordance with the manufacturer’s instructions. There is no additional washing requirement above what would normally be carried out. | How are you washing laundry? | [Cleaning (non-healthcare settings)](https://www.gov.uk/government/publications/covid-19-decontamination-in-non-healthcare-settings/covid-19-decontamination-in-non-healthcare-settings) |
| Waste | Waste does not need to be segregated unless an individual in the setting shows symptoms of or tests positive for COVID19. | How are you disposing of waste? | [Cleaning (non-healthcare settings)](https://www.gov.uk/government/publications/covid-19-decontamination-in-non-healthcare-settings/covid-19-decontamination-in-non-healthcare-settings) |
| Ventilation* | Setting any ventilation or air conditioning system that normally runs with a recirculation mode should to run on full outside air where possible. | Have you serviced or adjusted ventilation systems, if these are available? | [Hotels and guest accommodation 5.3](https://www.gov.uk/guidance/working-safely-during-coronavirus-covid-19/hotels-and-other-guest-accommodation#section-5-3)  [Places of worship 5](https://www.gov.uk/government/publications/covid-19-guidance-for-the-safe-use-of-places-of-worship-during-the-pandemic-from-4-july/covid-19-guidance-for-the-safe-use-of-places-of-worship-during-the-pandemic-from-4-july)  [Visitor economy 2.2](https://www.gov.uk/guidance/working-safely-during-coronavirus-covid-19/the-visitor-economy#shops-4-2) |
|  | Operating the ventilation system 24 hours a day. | Have you serviced or adjusted ventilation systems, if these are available? | [Visitor economy 2.2](https://www.gov.uk/guidance/working-safely-during-coronavirus-covid-19/the-visitor-economy#shops-4-2) |
|  | Increasing the frequency of filter changes. | Have you serviced or adjusted ventilation systems, if these are available? | [Visitor economy 2.2](https://www.gov.uk/guidance/working-safely-during-coronavirus-covid-19/the-visitor-economy#shops-4-2) |
|  | In the absence of known ventilation rates, a carbon dioxide sensor shall be used as a surrogate indicator to switch on additional mechanical ventilation or open windows. | Have you serviced or adjusted ventilation systems, if these are available? | [Visitor economy 2.2](https://www.gov.uk/guidance/working-safely-during-coronavirus-covid-19/the-visitor-economy#shops-4-2) |

*Further details available from [The Chartered Institution of Building Services Engineers guidance](https://www.cibse.org/coronavirus-covid-19/emerging-from-lockdown)

#### 6.2 Toilets

| **Key considerations** | **Potential mitigations** | **Suggested questions** | **Relevant guidance** |
| --- | --- | --- | --- |
| Guest and worker awareness | Using signs and posters to build awareness of good handwashing technique, the need to increase handwashing frequency and to avoid touching your face. | Have you used signs and posters to build awareness of handwashing technique and frequency? Please provide details. | [Performing arts 3.13](https://www.gov.uk/guidance/working-safely-during-coronavirus-covid-19/performing-arts#arts-3-13)  [Performing arts 5.3](https://www.gov.uk/guidance/working-safely-during-coronavirus-covid-19/performing-arts#arts-5-3) [Hotels and guest accommodation 5.3](https://www.gov.uk/guidance/working-safely-during-coronavirus-covid-19/hotels-and-other-guest-accommodation#section-5-3)  [Places of worship 5](https://www.gov.uk/government/publications/covid-19-guidance-for-the-safe-use-of-places-of-worship-during-the-pandemic-from-4-july/covid-19-guidance-for-the-safe-use-of-places-of-worship-during-the-pandemic-from-4-july)  [Restaurants, pubs, bars and takeaway services 2.4](https://www.gov.uk/guidance/working-safely-during-coronavirus-covid-19/restaurants-offering-takeaway-or-delivery#takeaways-2-4)  [Restaurants, pubs, bars and takeaway services 5.4](https://www.gov.uk/guidance/working-safely-during-coronavirus-covid-19/restaurants-offering-takeaway-or-delivery#takeaways-5-4)  [Food businesses](https://www.gov.uk/government/publications/covid-19-guidance-for-food-businesses/guidance-for-food-businesses-on-coronavirus-covid-19)  [Visitor economy 2.1](https://www.gov.uk/guidance/working-safely-during-coronavirus-covid-19/the-visitor-economy#shops-4-1)  [Visitor economy 5.3](https://www.gov.uk/guidance/working-safely-during-coronavirus-covid-19/the-visitor-economy#shops-5-3) |
|  | Consider the use of social distancing marking in areas where queues normally form, and the adoption of a limited entry approach. | Have you used social distancing markers and limited entry approaches around toilets? | [Food businesses](https://www.gov.uk/government/publications/covid-19-guidance-for-food-businesses/guidance-for-food-businesses-on-coronavirus-covid-19)  [Hotels and guest accommodation 5.3](https://www.gov.uk/guidance/working-safely-during-coronavirus-covid-19/hotels-and-other-guest-accommodation#section-5-3)  [Performing arts 3.13](https://www.gov.uk/guidance/working-safely-during-coronavirus-covid-19/performing-arts#arts-3-13)  [Places of worship 5](https://www.gov.uk/government/publications/covid-19-guidance-for-the-safe-use-of-places-of-worship-during-the-pandemic-from-4-july/covid-19-guidance-for-the-safe-use-of-places-of-worship-during-the-pandemic-from-4-july)  [Restaurants, pubs, bars and takeaway services 2.4](https://www.gov.uk/guidance/working-safely-during-coronavirus-covid-19/restaurants-offering-takeaway-or-delivery#takeaways-2-4)  [Visitor economy 2.1](https://www.gov.uk/guidance/working-safely-during-coronavirus-covid-19/the-visitor-economy#shops-4-1)  [Visitor economy 5.3](https://www.gov.uk/guidance/working-safely-during-coronavirus-covid-19/the-visitor-economy#shops-5-3) |
|  | Where toilets are shared, ensuring they are kept clean and clear of personal items. | Are shared toilets clear of personal items? | [Cleaning (non-healthcare settings)](https://www.gov.uk/government/publications/covid-19-decontamination-in-non-healthcare-settings/covid-19-decontamination-in-non-healthcare-settings)  [Hotels and guest accommodation 2.1](https://www.gov.uk/guidance/working-safely-during-coronavirus-covid-19/hotels-and-other-guest-accommodation#section-2-1)  [Visitor economy 2.1](https://www.gov.uk/guidance/working-safely-during-coronavirus-covid-19/the-visitor-economy#shops-4-1)  [Visitor economy 5.3](https://www.gov.uk/guidance/working-safely-during-coronavirus-covid-19/the-visitor-economy#shops-5-3) |
| Handwashing | Consider making hand sanitiser available on entry to toilets, and ensure suitable handwashing facilities and options for drying are available. | Is hand sanitiser available on entry to toilets? | [Hotels and guest accommodation 2.2](https://www.gov.uk/guidance/working-safely-during-coronavirus-covid-19/hotels-and-other-guest-accommodation#section-2-2)  [Performing arts 3.13](https://www.gov.uk/guidance/working-safely-during-coronavirus-covid-19/performing-arts#arts-3-13)  [Performing arts 5.3](https://www.gov.uk/guidance/working-safely-during-coronavirus-covid-19/performing-arts#arts-5-3)  [Places of worship 5](https://www.gov.uk/government/publications/covid-19-guidance-for-the-safe-use-of-places-of-worship-during-the-pandemic-from-4-july/covid-19-guidance-for-the-safe-use-of-places-of-worship-during-the-pandemic-from-4-july)  [Restaurants, pubs, bars and takeaway services 2.4](https://www.gov.uk/guidance/working-safely-during-coronavirus-covid-19/restaurants-offering-takeaway-or-delivery#takeaways-2-4)  [Cleaning (non-healthcare settings)](https://www.gov.uk/government/publications/covid-19-decontamination-in-non-healthcare-settings/covid-19-decontamination-in-non-healthcare-settings)  [Food businesses](https://www.gov.uk/government/publications/covid-19-guidance-for-food-businesses/guidance-for-food-businesses-on-coronavirus-covid-19)  [Visitor economy 2.1](https://www.gov.uk/guidance/working-safely-during-coronavirus-covid-19/the-visitor-economy#shops-4-1)  [Visitor economy 5.3](https://www.gov.uk/guidance/working-safely-during-coronavirus-covid-19/the-visitor-economy#shops-5-3) |
|  | Providing hand sanitiser in multiple locations in addition to washrooms. | Where, in addition to washrooms, is hand sanitiser available in your venue? | [Hotels and guest accommodation 5.3](https://www.gov.uk/guidance/working-safely-during-coronavirus-covid-19/hotels-and-other-guest-accommodation#section-5-3)  [Places of worship 5](https://www.gov.uk/government/publications/covid-19-guidance-for-the-safe-use-of-places-of-worship-during-the-pandemic-from-4-july/covid-19-guidance-for-the-safe-use-of-places-of-worship-during-the-pandemic-from-4-july)  [Restaurants, pubs, bars and takeaway services 5.4](https://www.gov.uk/guidance/working-safely-during-coronavirus-covid-19/restaurants-offering-takeaway-or-delivery#takeaways-5-4) |
|  | Providing hand drying facilities. | Are hand drying facilities provided? | [Cleaning (non-healthcare settings)](https://www.gov.uk/government/publications/covid-19-decontamination-in-non-healthcare-settings/covid-19-decontamination-in-non-healthcare-settings)  [Hotels and guest accommodation 5.3](https://www.gov.uk/guidance/working-safely-during-coronavirus-covid-19/hotels-and-other-guest-accommodation#section-5-3)  [Places of worship 5](https://www.gov.uk/government/publications/covid-19-guidance-for-the-safe-use-of-places-of-worship-during-the-pandemic-from-4-july/covid-19-guidance-for-the-safe-use-of-places-of-worship-during-the-pandemic-from-4-july)  [Visitor economy 5.3](https://www.gov.uk/guidance/working-safely-during-coronavirus-covid-19/the-visitor-economy#shops-5-3) |
| Cleaning | Setting clear use and cleaning guidance for toilets, with increased frequency of cleaning in line with usage. As a minimum, frequently touched surfaces should be wiped down twice a day, and one of these should be at the beginning or the end of the working day. Cleaning should be more frequent depending on the number of people using the space, whether they are entering and exiting the setting and access to handwashing and hand-sanitising facilities. Cleaning of frequently touched surfaces is particularly important in bathrooms and communal kitchens. | Please provide details of your cleaning guidance for toilets, including frequency of cleaning.  In addition, please state the number of facilities available at your venue. | [Performing arts 3.13](https://www.gov.uk/guidance/working-safely-during-coronavirus-covid-19/performing-arts#arts-3-13)  [Performing arts 5.3](https://www.gov.uk/guidance/working-safely-during-coronavirus-covid-19/performing-arts#arts-5-3)  [Places of worship 5](https://www.gov.uk/government/publications/covid-19-guidance-for-the-safe-use-of-places-of-worship-during-the-pandemic-from-4-july/covid-19-guidance-for-the-safe-use-of-places-of-worship-during-the-pandemic-from-4-july)  [Hotels and guest accommodation 2.1](https://www.gov.uk/guidance/working-safely-during-coronavirus-covid-19/hotels-and-other-guest-accommodation#section-2-1)  [Hotels and guest accommodation 5.3](https://www.gov.uk/guidance/working-safely-during-coronavirus-covid-19/hotels-and-other-guest-accommodation#section-5-3)  [Restaurants, pubs, bars and takeaway services 2.4](https://www.gov.uk/guidance/working-safely-during-coronavirus-covid-19/restaurants-offering-takeaway-or-delivery#takeaways-2-4)  [Restaurants, pubs, bars and takeaway services 5.4](https://www.gov.uk/guidance/working-safely-during-coronavirus-covid-19/restaurants-offering-takeaway-or-delivery#takeaways-5-4)  [Cleaning (non-healthcare settings)](https://www.gov.uk/government/publications/covid-19-decontamination-in-non-healthcare-settings/covid-19-decontamination-in-non-healthcare-settings)  [Food businesses](https://www.gov.uk/government/publications/covid-19-guidance-for-food-businesses/guidance-for-food-businesses-on-coronavirus-covid-19)  [Visitor economy 2.1](https://www.gov.uk/guidance/working-safely-during-coronavirus-covid-19/the-visitor-economy#shops-4-1)  [Visitor economy 5.3](https://www.gov.uk/guidance/working-safely-during-coronavirus-covid-19/the-visitor-economy#shops-5-3) |
|  | Keep the facilities well ventilated, for example by fixing doors open where appropriate. | How have you ensured facilities are ventilated? | [Performing arts 3.13](https://www.gov.uk/guidance/working-safely-during-coronavirus-covid-19/performing-arts#arts-3-13)  [Places of worship 5](https://www.gov.uk/government/publications/covid-19-guidance-for-the-safe-use-of-places-of-worship-during-the-pandemic-from-4-july/covid-19-guidance-for-the-safe-use-of-places-of-worship-during-the-pandemic-from-4-july)  [Hotels and guest accommodation 2.1](https://www.gov.uk/guidance/working-safely-during-coronavirus-covid-19/hotels-and-other-guest-accommodation#section-2-1)  [Hotels and guest accommodation 5.3](https://www.gov.uk/guidance/working-safely-during-coronavirus-covid-19/hotels-and-other-guest-accommodation#section-5-3)  [Restaurants, pubs, bars and takeaway services 2.4](https://www.gov.uk/guidance/working-safely-during-coronavirus-covid-19/restaurants-offering-takeaway-or-delivery#takeaways-2-4)  [Food businesses](https://www.gov.uk/government/publications/covid-19-guidance-for-food-businesses/guidance-for-food-businesses-on-coronavirus-covid-19)  [Visitor economy 2.1](https://www.gov.uk/guidance/working-safely-during-coronavirus-covid-19/the-visitor-economy#shops-4-1)  [Visitor economy 5.3](https://www.gov.uk/guidance/working-safely-during-coronavirus-covid-19/the-visitor-economy#shops-5-3) |
|  | Special care should be taken for cleaning of portable toilets and larger toilet blocks. Cleaning of frequently touched surfaces is particularly important in bathrooms and communal kitchens. | Do you have portable toilets or larger toilet blocks in your venue? Please provide details of how often these will be cleaned. | [Performing arts 3.13](https://www.gov.uk/guidance/working-safely-during-coronavirus-covid-19/performing-arts#arts-3-13)  [Places of worship 5](https://www.gov.uk/government/publications/covid-19-guidance-for-the-safe-use-of-places-of-worship-during-the-pandemic-from-4-july/covid-19-guidance-for-the-safe-use-of-places-of-worship-during-the-pandemic-from-4-july)  [Hotels and guest accommodation 5.3](https://www.gov.uk/guidance/working-safely-during-coronavirus-covid-19/hotels-and-other-guest-accommodation#section-5-3)  [Restaurants, pubs, bars and takeaway services 2.4](https://www.gov.uk/guidance/working-safely-during-coronavirus-covid-19/restaurants-offering-takeaway-or-delivery#takeaways-2-4)  [Restaurants, pubs, bars and takeaway services 5.4](https://www.gov.uk/guidance/working-safely-during-coronavirus-covid-19/restaurants-offering-takeaway-or-delivery#takeaways-5-4)  [Cleaning (non-healthcare settings)](https://www.gov.uk/government/publications/covid-19-decontamination-in-non-healthcare-settings/covid-19-decontamination-in-non-healthcare-settings)  [Food businesses](https://www.gov.uk/government/publications/covid-19-guidance-for-food-businesses/guidance-for-food-businesses-on-coronavirus-covid-19)  [Visitor economy 5.3](https://www.gov.uk/guidance/working-safely-during-coronavirus-covid-19/the-visitor-economy#shops-5-3) |
|  | Putting up a visible cleaning schedule and keeping it up to date. | Do you have a visible cleaning schedule near toilets? | [Performing arts 3.13](https://www.gov.uk/guidance/working-safely-during-coronavirus-covid-19/performing-arts#arts-3-13)  [Places of worship 5](https://www.gov.uk/government/publications/covid-19-guidance-for-the-safe-use-of-places-of-worship-during-the-pandemic-from-4-july/covid-19-guidance-for-the-safe-use-of-places-of-worship-during-the-pandemic-from-4-july)  [Hotels and guest accommodation 5.3](https://www.gov.uk/guidance/working-safely-during-coronavirus-covid-19/hotels-and-other-guest-accommodation#section-5-3)  [Restaurants, pubs, bars and takeaway services 2.4](https://www.gov.uk/guidance/working-safely-during-coronavirus-covid-19/restaurants-offering-takeaway-or-delivery#takeaways-2-4)  [Food businesses](https://www.gov.uk/government/publications/covid-19-guidance-for-food-businesses/guidance-for-food-businesses-on-coronavirus-covid-19)  [Visitor economy 5.3](https://www.gov.uk/guidance/working-safely-during-coronavirus-covid-19/the-visitor-economy#shops-5-3) |
|  | Providing more waste facilities and more frequent rubbish collection. | How often will rubbish collection be undertaken? | [Performing arts 3.13](https://www.gov.uk/guidance/working-safely-during-coronavirus-covid-19/performing-arts#arts-3-13)  [Performing arts 5.3](https://www.gov.uk/guidance/working-safely-during-coronavirus-covid-19/performing-arts#arts-5-3)  [Places of worship 5](https://www.gov.uk/government/publications/covid-19-guidance-for-the-safe-use-of-places-of-worship-during-the-pandemic-from-4-july/covid-19-guidance-for-the-safe-use-of-places-of-worship-during-the-pandemic-from-4-july)  [Hotels and guest accommodation 5.3](https://www.gov.uk/guidance/working-safely-during-coronavirus-covid-19/hotels-and-other-guest-accommodation#section-5-3)  [Restaurants, pubs, bars and takeaway services 2.4](https://www.gov.uk/guidance/working-safely-during-coronavirus-covid-19/restaurants-offering-takeaway-or-delivery#takeaways-2-4)  [Restaurants, pubs, bars and takeaway services 5.4](https://www.gov.uk/guidance/working-safely-during-coronavirus-covid-19/restaurants-offering-takeaway-or-delivery#takeaways-5-4)  [Food businesses](https://www.gov.uk/government/publications/covid-19-guidance-for-food-businesses/guidance-for-food-businesses-on-coronavirus-covid-19)  [Visitor economy 5.3](https://www.gov.uk/guidance/working-safely-during-coronavirus-covid-19/the-visitor-economy#shops-5-3) |
| Patterns of use | Considering the likely patterns of use during a performance, for example during intervals and modifying to avoid the creation of pinch points. | How have you considered the patterns of use of toilet facilities? | [Performing arts 3.13](https://www.gov.uk/guidance/working-safely-during-coronavirus-covid-19/performing-arts#arts-3-13) |
|  | Encouraging staggered use of washroom facilities wherever possible. | Have you encouraged staggered use of toilet facilities? | [Hotels and guest accommodation 5.3](https://www.gov.uk/guidance/working-safely-during-coronavirus-covid-19/hotels-and-other-guest-accommodation#section-5-3) |

#### 6.3 Shower facilities

Only applies if shower facilities are available as part of the event.

| **Key considerations** | **Potential mitigations** | **Suggested questions** | **Relevant guidance** |
| --- | --- | --- | --- |
| Closing indoor facilities | Either shutting shared shower facilities or assigning them to one household group or support bubble, (i.e. making them private), or running a reservation and clean process (whereby one household can exclusively book the shared facilities for a fixed time, and the facilities are cleaned thoroughly between reservations). | How have you modified the processes for shared shower facilities? | [Hotels and guest accommodation 2.1](https://www.gov.uk/guidance/working-safely-during-coronavirus-covid-19/hotels-and-other-guest-accommodation#section-2-1)  [Hotels and guest accommodation 5.3](https://www.gov.uk/guidance/working-safely-during-coronavirus-covid-19/hotels-and-other-guest-accommodation#section-5-3) |
| Attendee awareness | Making information available to guests on the increased risk of using shared facilities in accommodation. | Have you provided information on the increased risk of using shared facilities? | [Hotels and guest accommodation 2.1](https://www.gov.uk/guidance/working-safely-during-coronavirus-covid-19/hotels-and-other-guest-accommodation#section-2-1)  [Hotels and guest accommodation 5.3](https://www.gov.uk/guidance/working-safely-during-coronavirus-covid-19/hotels-and-other-guest-accommodation#section-5-3) |
| Cleaning practices | Setting clear use and cleaning guidance for showers, lockers and changing rooms to ensure they are kept clean and clear of personal items. | Please outline cleaning procedures and frequency for all shared shower facilities. | [Hotels and guest accommodation 5.3](https://www.gov.uk/guidance/working-safely-during-coronavirus-covid-19/hotels-and-other-guest-accommodation#section-5-3)  [Performing arts 4.6](https://www.gov.uk/guidance/working-safely-during-coronavirus-covid-19/performing-arts#arts-4-6) |
|  | Introducing enhanced cleaning of these facilities regularly during the day and at the end of the day. | Please outline cleaning procedures and frequency for all shared shower facilities. | [Hotels and guest accommodation 5.3](https://www.gov.uk/guidance/working-safely-during-coronavirus-covid-19/hotels-and-other-guest-accommodation#section-5-3)  [Performing arts 4.6](https://www.gov.uk/guidance/working-safely-during-coronavirus-covid-19/performing-arts#arts-4-6) |
|  | Where possible assigning one group of washing and showering facilities to one household group. | Have you assigned assigning one group of washing and showering facilities to one household group? | [Hotels and guest accommodation 5.3](https://www.gov.uk/guidance/working-safely-during-coronavirus-covid-19/hotels-and-other-guest-accommodation#section-5-3) |
|  | Considering introducing a system of staggered entry and booked timeslots for using shower facilities. | Have you introduced a system of staggered entry or booked timeslots for shower facilities? | [Hotels and guest accommodation 5.3](https://www.gov.uk/guidance/working-safely-during-coronavirus-covid-19/hotels-and-other-guest-accommodation#section-5-3) |

#### 6.4 Cleaning following suspected or confirmed COVID cases

| **Key considerations** | **Potential mitigations** | **Suggested questions** | **Relevant guidance** |
| --- | --- | --- | --- |
| Cleaning and disinfection | Public areas where a symptomatic person has passed through and spent minimal time but which are not visibly contaminated with body fluids, such as corridors, can be cleaned thoroughly as normal. | Please outline your cleaning procedures following suspected or confirmed COVID cases. | [Cleaning (non-healthcare settings)](https://www.gov.uk/government/publications/covid-19-decontamination-in-non-healthcare-settings/covid-19-decontamination-in-non-healthcare-settings) |
|  | All surfaces that the symptomatic person has come into contact with should be cleaned and disinfected, including all potentially contaminated and frequently touched areas such as bathrooms, door handles, telephones, grab rails in corridors and stairwells. | Please outline your cleaning procedures following suspected or confirmed COVID cases. | [Cleaning (non-healthcare settings)](https://www.gov.uk/government/publications/covid-19-decontamination-in-non-healthcare-settings/covid-19-decontamination-in-non-healthcare-settings) |
|  | Keep people away from the area. Use a spill-kit if available, using the PPE in the kit or PPE provided by your employer/organisation and following the instructions provided with the spill-kit. If no spill-kit is available, place paper towels/roll onto the spill, and seek further advice from emergency services when they arrive. | Please outline your cleaning procedures following suspected or confirmed COVID cases. | [Guidance for first responders](https://www.gov.uk/government/publications/novel-coronavirus-2019-ncov-interim-guidance-for-first-responders/interim-guidance-for-first-responders-and-others-in-close-contact-with-symptomatic-people-with-potential-2019-ncov) |
| Laundry | Wash items in accordance with the manufacturer’s instructions. Use the warmest water setting and dry items completely. Dirty laundry that has been in contact with an unwell person can be washed with other people’s items. To minimise the possibility of dispersing virus through the air, do not shake dirty laundry prior to washing. | Please outline your cleaning procedures following suspected or confirmed COVID cases. | [Cleaning (non-healthcare settings)](https://www.gov.uk/government/publications/covid-19-decontamination-in-non-healthcare-settings/covid-19-decontamination-in-non-healthcare-settings) |

### 7 COVID risk domain: accommodation

This section applies only if accommodation is provided as part of the event.

Please note that guidance is specific to each accommodation type. General principles are outlined below, and further detail is available in the [hotels and other guest accommodation guidance](https://www.gov.uk/guidance/working-safely-during-coronavirus-covid-19/hotels-and-other-guest-accommodation).

In addition, further guidance from other bodies for the visitor economy is linked in 2.3.1 of the above guidance, and business events guidance in 2.3.2. This may be useful to consider when assessing risk; however, given the location-specific nature of this guidance, it has not been included below.

#### 7.1 Employer-provided accommodation

Only applies where the employer is providing accommodation for staff during the event.

| **Key considerations** | **Potential mitigations** | **Suggested questions** | **Relevant guidance** |
| --- | --- | --- | --- |
| Occupancy | Secure single occupancy accommodation for workers. If this is not possible, occupancy in each shared space should be as low as possible | How many workers share a single space? | [Food businesses](https://www.gov.uk/government/publications/covid-19-guidance-for-food-businesses/guidance-for-food-businesses-on-coronavirus-covid-19) |
|  | Organise workers in shared residential accommodation to be within the same cohort in the workplace where possible. | Are shared residential workers in the same cohort in the workplace? | [Food businesses](https://www.gov.uk/government/publications/covid-19-guidance-for-food-businesses/guidance-for-food-businesses-on-coronavirus-covid-19) |
|  | Make arrangements for back-up single occupancy accommodation to allow workers who become unwell to self-isolate | Do you have back-up single occupancy accommodation in case workers become unwell? | [Food businesses](https://www.gov.uk/government/publications/covid-19-guidance-for-food-businesses/guidance-for-food-businesses-on-coronavirus-covid-19) |
| Social distancing | Schedule access to shared spaces such as kitchens and living areas in the provided accommodation to limit crowding and promote social distancing | Have you scheduled access to shared spaces e.g. kitchens and living areas? | [Food businesses](https://www.gov.uk/government/publications/covid-19-guidance-for-food-businesses/guidance-for-food-businesses-on-coronavirus-covid-19) |
|  | Where workers are required to stay away from their home, centrally logging the stay and confirming that any overnight accommodation meets social distancing guidelines. | Please provide details of how information of workers is logged. | [Visitor economy 7.2](https://www.gov.uk/guidance/working-safely-during-coronavirus-covid-19/the-visitor-economy#shops-7-2) |

#### 7.2 Modification to accommodation

| **Key considerations** | **Potential mitigations** | **Suggested questions** | **Relevant guidance** |
| --- | --- | --- | --- |
| Modification of communal areas | Taking measures to make reception areas safer, with increased cleaning, keeping the activity time as short as possible and considering the addition of screens between guests and staff. | How have you taken steps to ensure reception areas are safer? | [Hotels and guest accommodation 2.1](https://www.gov.uk/guidance/working-safely-during-coronavirus-covid-19/hotels-and-other-guest-accommodation#section-2-1)  [Visitor economy 4.3](https://www.gov.uk/guidance/working-safely-during-coronavirus-covid-19/the-visitor-economy#shops-3-3) |
|  | Considering minimising lift usage from reception. | Have you minimised lift usage from reception? | [Hotels and guest accommodation 2.1](https://www.gov.uk/guidance/working-safely-during-coronavirus-covid-19/hotels-and-other-guest-accommodation#section-2-1) |
|  | Closing dormitories and shared indoor facilities, including kitchens. | Have you closed dormitories and shared indoor facilities, including kitchens? | [Hotels and guest accommodation 2.1](https://www.gov.uk/guidance/working-safely-during-coronavirus-covid-19/hotels-and-other-guest-accommodation#section-2-1) |
|  | Considering removal of items that are likely to be regularly touched by lots of different people, for example shared newspapers. | Have you removed items likely to be touched by different people e.g. shared newspapers? | [Hotels and guest accommodation 5.2](https://www.gov.uk/guidance/working-safely-during-coronavirus-covid-19/hotels-and-other-guest-accommodation#section-5-2) |
| Check in procedures | Taking measures to ensure the handover of keys to property can be done in a socially distanced way, ensuring that keys are cleaned. | How have you ensured that handover of keys to property can be done in a socially distanced way? | [Hotels and guest accommodation 2.1](https://www.gov.uk/guidance/working-safely-during-coronavirus-covid-19/hotels-and-other-guest-accommodation#section-2-1) |
|  | Taking measures to avoid crowded reception areas, such as staggering check-in and check-out times or placing markers on the floor to maintain social distancing. | How have you ensured that reception areas will not be crowded? | [Hotels and guest accommodation 2.2](https://www.gov.uk/guidance/working-safely-during-coronavirus-covid-19/hotels-and-other-guest-accommodation#section-2-2) |
|  | Encouraging contactless payments or pre-payments for rooms as part of the online booking, where possible, to limit cash payments for bills. | Have you encouraged contactless or pre-payments for rooms? | [Hotels and guest accommodation 2.2](https://www.gov.uk/guidance/working-safely-during-coronavirus-covid-19/hotels-and-other-guest-accommodation#section-2-2) |
|  | Informing guests about preventative measures being taken and other services they may require. | What preventative measures are you informing guests about? | [Hotels and guest accommodation 2.3](https://www.gov.uk/guidance/working-safely-during-coronavirus-covid-19/hotels-and-other-guest-accommodation#section-2-3) |
| Providing guidance on COVID-symptomatic guests | Accommodation providers should consider how best to inform guests about their policy for COVID-symptomatic guests, for example during the booking or check-in process. | How are you providing information on your COVID-symptomatic policy during check-in to accommodation? | [Hotels and guest accommodation 5.2](https://www.gov.uk/guidance/working-safely-during-coronavirus-covid-19/hotels-and-other-guest-accommodation#section-5-2) |

#### 7.3 Accessing accommodation

| **Key considerations** | **Potential mitigations** | **Suggested questions** | **Relevant guidance** |
| --- | --- | --- | --- |
| Ease of access, crowd control | See “crowd control” and “ingress and egress” sections above. | As per relevant sections. | As per relevant sections. |

#### 7.4 Managing contacts

| **Key considerations** | **Potential mitigations** | **Suggested questions** | **Relevant guidance** |
| --- | --- | --- | --- |
| Minimise the contact resulting from visits | Informing guests and contractors of guidance about visiting the premises prior to and at the point of arrival (including information on websites, on booking forms and in entrance ways). | How have you provided information to guests and contractors prior to, and on the point of, arrival? | [Hotels and guest accommodation 2.2](https://www.gov.uk/guidance/working-safely-during-coronavirus-covid-19/hotels-and-other-guest-accommodation#section-2-2)  [Visitor economy 2.2](https://www.gov.uk/guidance/working-safely-during-coronavirus-covid-19/the-visitor-economy#shops-4-2) |
|  | Working with neighbouring businesses and local authorities to consider how to spread the number of people arriving throughout the day. | How have you spread the number of people arriving throughout the day? | [Hotels and guest accommodation 2.2](https://www.gov.uk/guidance/working-safely-during-coronavirus-covid-19/hotels-and-other-guest-accommodation#section-2-2) |
|  | Making staff accessible to guests via phone, emails and guest apps. | Have you made staff accessible to guests in a non-face to face way? | [Hotels and guest accommodation 2.2](https://www.gov.uk/guidance/working-safely-during-coronavirus-covid-19/hotels-and-other-guest-accommodation#section-2-2) |

#### 7.5 Providing information to guests

| **Key considerations** | **Potential mitigations** | **Suggested questions** | **Relevant guidance** |
| --- | --- | --- | --- |
| Providing information to guests | See “information for guests and workers” section above. | As per relevant section. | As per relevant section. |

#### 7.6 Housekeeping

| **Key considerations** | **Potential mitigations** | **Suggested questions** | **Relevant guidance** |
| --- | --- | --- | --- |
| Maintaining cleanliness | Consider the ability to perform housekeeping, whilst maintaining social distancing. | How will you maintain social distancing whilst housekeeping? | [Hotels and guest accommodation 5.2](https://www.gov.uk/guidance/working-safely-during-coronavirus-covid-19/hotels-and-other-guest-accommodation#section-5-2) |
|  | When cleaning a room, focus on cleaning of all hand contact surfaces in rooms. | Please detail cleaning procedures as part of housekeeping. | [Hotels and guest accommodation 5.2](https://www.gov.uk/guidance/working-safely-during-coronavirus-covid-19/hotels-and-other-guest-accommodation#section-5-2) |
|  | Considering removal of items from the room that are not likely to be needed by guests. | Please detail cleaning procedures as part of housekeeping. | [Hotels and guest accommodation 5.2](https://www.gov.uk/guidance/working-safely-during-coronavirus-covid-19/hotels-and-other-guest-accommodation#section-5-2) |
|  | Glasses and crockery should be removed and washed between guests. | Please detail cleaning procedures as part of housekeeping. | [Hotels and guest accommodation 5.2](https://www.gov.uk/guidance/working-safely-during-coronavirus-covid-19/hotels-and-other-guest-accommodation#section-5-2) |
|  | Towels and linens should be washed in accordance with washing instructions. The frequency of the cycle of cleaning and in-room services should be reviewed to take into account different lengths of stay. | Please detail cleaning procedures as part of housekeeping. | [Hotels and guest accommodation 5.2](https://www.gov.uk/guidance/working-safely-during-coronavirus-covid-19/hotels-and-other-guest-accommodation#section-5-2) |
|  | Making a checklist of all hand contact services to be cleaned when each guest vacates. | Have you provided a checklist of all areas to be cleaned when guests vacate? | [Hotels and guest accommodation 2.1](https://www.gov.uk/guidance/working-safely-during-coronavirus-covid-19/hotels-and-other-guest-accommodation#section-2-1) |
| Increased cleaning | Considering increased surface cleaning for confined accommodation such as tents or caravans and leaving longer periods between usage by different guests. | Have you increased cleaning or periods between usage of accommodation? | [Hotels and guest accommodation 5.2](https://www.gov.uk/guidance/working-safely-during-coronavirus-covid-19/hotels-and-other-guest-accommodation#section-5-2) |

#### 7.7 Food and drink facilities in accommodation

Only applies if food and drinks are sold as part of accommodation.

| **Key considerations** | **Potential mitigations** | **Suggested questions** | **Relevant guidance** |
| --- | --- | --- | --- |
| Selling food and drink | See “food and drink facilities” section below. | As per relevant section. | As per relevant section. |
| Room service | Asking customers to order room service over the telephone if they are staying in accommodation. | Have you requested customers in accommodation to order room service? | [Hotels and guest accommodation 2.2](https://www.gov.uk/guidance/working-safely-during-coronavirus-covid-19/hotels-and-other-guest-accommodation#section-2-2) |

#### 7.8 Shared facilities

Only applies if shared kitchen or toilet facilities are available as part of accommodation.

| **Key considerations** | **Potential mitigations** | **Suggested questions** | **Relevant guidance** |
| --- | --- | --- | --- |
| Closing indoor kitchen facilities | Either shutting shared kitchen facilities or assigning them to one household group or support bubble, (i.e. making them private), or running a reservation and clean process (whereby one household can exclusively book the shared facilities for a fixed time, and the facilities are cleaned thoroughly between reservations). | How have you altered the use of indoor shared kitchen facilities? | [Hotels and guest accommodation 5.3](https://www.gov.uk/guidance/working-safely-during-coronavirus-covid-19/hotels-and-other-guest-accommodation#section-5-3) |
| Signposting of information | Making information available to guests on the increased risk of using shared facilities in accommodation. | Have you informed guests about the increased risk of using shared facilities in accommodation? | [Hotels and guest accommodation 2.1](https://www.gov.uk/guidance/working-safely-during-coronavirus-covid-19/hotels-and-other-guest-accommodation#section-2-1) |

### 8 COVID risk domain: Food and drink facilities

This section applies only if food and drink facilities (but not those which are self-catered – see accommodation above for these) are available as part of the event.

#### 8.1 Minimising contacts and maintaining social distancing

| **Key considerations** | **Potential mitigations** | **Suggested questions** | **Relevant guidance** |
| --- | --- | --- | --- |
| Minimising contact | Ensuring that any bar or dining area is only opened in a way compliant with UK government guidance on the hospitality sector. | Have you consulted the UK government guidance on the hospitality sector? | [Hotels and guest accommodation 2.1](https://www.gov.uk/guidance/working-safely-during-coronavirus-covid-19/hotels-and-other-guest-accommodation#section-2-1) |
|  | Maintaining social distancing when taking orders from customers. | How are you maintaining social distancing when taking orders from customers? | [Hotels and guest accommodation 2.2](https://www.gov.uk/guidance/working-safely-during-coronavirus-covid-19/hotels-and-other-guest-accommodation#section-2-2) |
|  | Using social distance markings to remind customers to maintain social distancing. | Are you using social distancing markers to remind customers? | [Hotels and guest accommodation 2.2](https://www.gov.uk/guidance/working-safely-during-coronavirus-covid-19/hotels-and-other-guest-accommodation#section-2-2)  [Restaurants, pubs, bars and takeaway services 2.2](https://www.gov.uk/guidance/working-safely-during-coronavirus-covid-19/restaurants-offering-takeaway-or-delivery#takeaways-2-2) |
|  | Minimising customer self-service of food, cutlery and condiments to reduce risk of transmission. | Have you removed customer self-service of food, cutlery and condiments? | [Hotels and guest accommodation 2.2](https://www.gov.uk/guidance/working-safely-during-coronavirus-covid-19/hotels-and-other-guest-accommodation#section-2-2)  [Restaurants, pubs, bars and takeaway services 2.2](https://www.gov.uk/guidance/working-safely-during-coronavirus-covid-19/restaurants-offering-takeaway-or-delivery#takeaways-2-2)  [Food businesses](https://www.gov.uk/government/publications/covid-19-guidance-for-food-businesses/guidance-for-food-businesses-on-coronavirus-covid-19) |
|  | Encouraging contactless payments where possible and adjusting location of card readers to social distancing guidelines. | Have you encouraged contactless payments? | [Hotels and guest accommodation 2.2](https://www.gov.uk/guidance/working-safely-during-coronavirus-covid-19/hotels-and-other-guest-accommodation#section-2-2)  [Restaurants, pubs, bars and takeaway services 2.2](https://www.gov.uk/guidance/working-safely-during-coronavirus-covid-19/restaurants-offering-takeaway-or-delivery#takeaways-2-2)  [Food businesses](https://www.gov.uk/government/publications/covid-19-guidance-for-food-businesses/guidance-for-food-businesses-on-coronavirus-covid-19) |
|  | Providing only disposable condiments or cleaning non-disposable condiment containers after each use. | Have you removed customer self-service of food, cutlery and condiments? | [Hotels and guest accommodation 2.2](https://www.gov.uk/guidance/working-safely-during-coronavirus-covid-19/hotels-and-other-guest-accommodation#section-2-2)  [Restaurants, pubs, bars and takeaway services 2.2](https://www.gov.uk/guidance/working-safely-during-coronavirus-covid-19/restaurants-offering-takeaway-or-delivery#takeaways-2-2) |
|  | Reducing the number of surfaces touched by both staff and customers. For example, asking customers to remain at a table where possible, or to not lean on counters when collecting takeaways. | How are you reducing the number of surfaces touched by staff and customers? | [Hotels and guest accommodation 2.2](https://www.gov.uk/guidance/working-safely-during-coronavirus-covid-19/hotels-and-other-guest-accommodation#section-2-2)  [Restaurants, pubs, bars and takeaway services 2.2](https://www.gov.uk/guidance/working-safely-during-coronavirus-covid-19/restaurants-offering-takeaway-or-delivery#takeaways-2-2)  [Food businesses](https://www.gov.uk/government/publications/covid-19-guidance-for-food-businesses/guidance-for-food-businesses-on-coronavirus-covid-19) |
|  | Ensuring all outdoor areas, with particular regard to covered areas, have sufficient ventilation. | Have you ensured all outdoor areas have sufficient ventilation? | [Hotels and guest accommodation 2.2](https://www.gov.uk/guidance/working-safely-during-coronavirus-covid-19/hotels-and-other-guest-accommodation#section-2-2)  [Restaurants, pubs, bars and takeaway services 2.2](https://www.gov.uk/guidance/working-safely-during-coronavirus-covid-19/restaurants-offering-takeaway-or-delivery#takeaways-2-2)  [Visitor economy 2.2](https://www.gov.uk/guidance/working-safely-during-coronavirus-covid-19/the-visitor-economy#shops-4-2) |
|  | Adjusting service approaches to minimise staff contact with customers. | How are you maintaining social distancing when taking orders from customers? | [Hotels and guest accommodation 2.2](https://www.gov.uk/guidance/working-safely-during-coronavirus-covid-19/hotels-and-other-guest-accommodation#section-2-2) |
|  | Adjusting processes to prevent customers from congregating at points of service. | How are you minimising queueing of customers? | [Hotels and guest accommodation 2.2](https://www.gov.uk/guidance/working-safely-during-coronavirus-covid-19/hotels-and-other-guest-accommodation#section-2-2) |
|  | Minimising contact between front of house workers and customers at points of service where appropriate. | How are you maintaining social distancing when taking orders from customers? | [Hotels and guest accommodation 2.2](https://www.gov.uk/guidance/working-safely-during-coronavirus-covid-19/hotels-and-other-guest-accommodation#section-2-2)  [Restaurants, pubs, bars and takeaway services 2.2](https://www.gov.uk/guidance/working-safely-during-coronavirus-covid-19/restaurants-offering-takeaway-or-delivery#takeaways-2-2) |
|  | Ventilation into the building should be optimised to ensure the maximum fresh air supply is provided to all areas of the facility wherever possible. | Have you ensured all indoor areas have sufficient ventilation? | [Restaurants, pubs, bars and takeaway services 2.3](https://www.gov.uk/guidance/working-safely-during-coronavirus-covid-19/restaurants-offering-takeaway-or-delivery#takeaways-2-3) |
|  | Ensure staff wear face coverings in hospitality settings. | Have you ensured all staff are wearing face coverings? | [Restaurants, pubs, bars and takeaway services](https://www.gov.uk/guidance/working-safely-during-coronavirus-covid-19/restaurants-offering-takeaway-or-delivery)  [Visitor economy](https://www.gov.uk/guidance/working-safely-during-coronavirus-covid-19/the-visitor-economy) |
|  | Let customers know that by law they can only visit in groups of up to 6 people (unless they are visiting as a household or support bubble which is larger than 6). | Have you ensured that all groups are informed that they can only visit in groups of up to 6? | [Restaurants, pubs, bars and takeaway services](https://www.gov.uk/guidance/working-safely-during-coronavirus-covid-19/restaurants-offering-takeaway-or-delivery)  [Visitor economy](https://www.gov.uk/guidance/working-safely-during-coronavirus-covid-19/the-visitor-economy) |
|  | Lower music and other background noise. | Have you lowered music and other background noise? | [Restaurants, pubs, bars and takeaway services](https://www.gov.uk/guidance/working-safely-during-coronavirus-covid-19/restaurants-offering-takeaway-or-delivery)  [Visitor economy](https://www.gov.uk/guidance/working-safely-during-coronavirus-covid-19/the-visitor-economy) |
|  | Ensure customers wear face coverings in retail and hospitality settings. | How will you ensure that customers wear face coverings? | [Restaurants, pubs, bars and takeaway services](https://www.gov.uk/guidance/working-safely-during-coronavirus-covid-19/restaurants-offering-takeaway-or-delivery)  [Visitor economy](https://www.gov.uk/guidance/working-safely-during-coronavirus-covid-19/the-visitor-economy) |

#### 8.2 Cleaning kitchen and food service areas

| **Key considerations** | **Potential mitigations** | **Suggested questions** | **Relevant guidance** |
| --- | --- | --- | --- |
| Maintaining cleanliness | Recognising that cleaning measures are already stringent in kitchen areas, consider the need for additional cleaning measures. | Have you considered the need for additional cleaning in kitchen areas? | [Hotels and guest accommodation 5.2](https://www.gov.uk/guidance/working-safely-during-coronavirus-covid-19/hotels-and-other-guest-accommodation#section-5-2)  [Restaurants, pubs, bars and takeaway services 5.3](https://www.gov.uk/guidance/working-safely-during-coronavirus-covid-19/restaurants-offering-takeaway-or-delivery#takeaways-5-3)  [Food businesses](https://www.gov.uk/government/publications/covid-19-guidance-for-food-businesses/guidance-for-food-businesses-on-coronavirus-covid-19) |
|  | Having bins for the collection of used towels and staff overalls. | Have you provided bins for the collection of used towels and staff overalls? | [Hotels and guest accommodation 5.2](https://www.gov.uk/guidance/working-safely-during-coronavirus-covid-19/hotels-and-other-guest-accommodation#section-5-2)  [Restaurants, pubs, bars and takeaway services 5.3](https://www.gov.uk/guidance/working-safely-during-coronavirus-covid-19/restaurants-offering-takeaway-or-delivery#takeaways-5-3) |
|  | Asking workers to wash hands before handling plates and takeaway boxes. | Have you asked workers to wash their hands before handling plates and takeaway boxes? | [Hotels and guest accommodation 5.2](https://www.gov.uk/guidance/working-safely-during-coronavirus-covid-19/hotels-and-other-guest-accommodation#section-5-2)  [Restaurants, pubs, bars and takeaway services 5.3](https://www.gov.uk/guidance/working-safely-during-coronavirus-covid-19/restaurants-offering-takeaway-or-delivery#takeaways-5-3)  [Food businesses](https://www.gov.uk/government/publications/covid-19-guidance-for-food-businesses/guidance-for-food-businesses-on-coronavirus-covid-19) |
|  | Continuing high frequency of hand washing throughout the day. | How have you encouraged hand-washing for workers? | [Hotels and guest accommodation 5.2](https://www.gov.uk/guidance/working-safely-during-coronavirus-covid-19/hotels-and-other-guest-accommodation#section-5-2)  [Restaurants, pubs, bars and takeaway services 5.3](https://www.gov.uk/guidance/working-safely-during-coronavirus-covid-19/restaurants-offering-takeaway-or-delivery#takeaways-5-3)  [Food businesses](https://www.gov.uk/government/publications/covid-19-guidance-for-food-businesses/guidance-for-food-businesses-on-coronavirus-covid-19) |
|  | Washing hands after handling customer items and before moving onto another task. For example, after collecting used plates for cleaning and before serving food to another table. | How have you encouraged hand-washing for workers? | [Restaurants, pubs, bars and takeaway services 5.4](https://www.gov.uk/guidance/working-safely-during-coronavirus-covid-19/restaurants-offering-takeaway-or-delivery#takeaways-5-4)  [Food businesses](https://www.gov.uk/government/publications/covid-19-guidance-for-food-businesses/guidance-for-food-businesses-on-coronavirus-covid-19) |

#### 8.3 Seating facilities

| **Key considerations** | **Potential mitigations** | **Suggested questions** | **Relevant guidance** |
| --- | --- | --- | --- |
| Supporting social distancing | Reconfiguring indoor and outdoor seating and tables to maintain social distancing guidelines. | How have you reconfigured seating facilities to maintain social distancing? | [Restaurants, pubs, bars and takeaway services 2.1](https://www.gov.uk/guidance/working-safely-during-coronavirus-covid-19/restaurants-offering-takeaway-or-delivery#takeaways-2-1) |
|  | Ensure all customers remain seated. | How have you ensured that customers remain seated to eat and drink at the venue? | [Restaurants, pubs, bars and takeaway services](https://www.gov.uk/guidance/working-safely-during-coronavirus-covid-19/restaurants-offering-takeaway-or-delivery)  [Visitor economy](https://www.gov.uk/guidance/working-safely-during-coronavirus-covid-19/the-visitor-economy) |
|  | If you sell alcohol, provide table service only. | If you sell alcohol, have you ensured that you only provide table service? | [Restaurants, pubs, bars and takeaway services](https://www.gov.uk/guidance/working-safely-during-coronavirus-covid-19/restaurants-offering-takeaway-or-delivery)  [Visitor economy](https://www.gov.uk/guidance/working-safely-during-coronavirus-covid-19/the-visitor-economy) |

#### 8.4 Other restrictions

| **Key considerations** | **Potential mitigations** | **Suggested questions** | **Relevant guidance** |
| --- | --- | --- | --- |
| Time restrictions | Businesses that sell food or drink (including cafes, bars, pubs and restaurants) must close services between 10pm - 5am. | Are you closing food and drink services between 10pm and 5am, if appropriate? | [Community 3](https://www.gov.uk/government/publications/covid-19-guidance-for-the-safe-use-of-multi-purpose-community-facilities/covid-19-guidance-for-the-safe-use-of-multi-purpose-community-facilities)  [Restaurants, pubs, bars and takeaway services](https://www.gov.uk/guidance/working-safely-during-coronavirus-covid-19/restaurants-offering-takeaway-or-delivery)  [Visitor economy](https://www.gov.uk/guidance/working-safely-during-coronavirus-covid-19/the-visitor-economy) |
| Preventing dancing | Preventing customers from dancing on the premises, except for a couple celebrating their wedding or civil partnership. This is required by law under the COVID-secure regulations. | Have you informed attendees that dancing is not allowed on the premises by law? | [Restaurants, pubs, bars and takeaway services 2.1](https://www.gov.uk/guidance/working-safely-during-coronavirus-covid-19/restaurants-offering-takeaway-or-delivery) |

### 9 COVID risk domain: Managing deliveries

This section applies only if there are on-site deliveries of goods and merchandise, e.g. for food and drink or retail facilities.

#### 9.1 Minimising transmission via goods and services

| **Key considerations** | **Potential mitigations** | **Suggested questions** | **Relevant guidance** |
| --- | --- | --- | --- |
| Minimising transmission via goods | Cleaning procedures for goods and merchandise entering the site. | Please outline cleaning procedures for goods and merchandise entering the site. | [Hotels and guest accommodation 5.4](https://www.gov.uk/guidance/working-safely-during-coronavirus-covid-19/hotels-and-other-guest-accommodation#section-5-4)  [Restaurants, pubs, bars and takeaway services 5.6](https://www.gov.uk/guidance/working-safely-during-coronavirus-covid-19/restaurants-offering-takeaway-or-delivery#takeaways-5-6)  [Visitor economy 5.4](https://www.gov.uk/guidance/working-safely-during-coronavirus-covid-19/the-visitor-economy#shops-5-4)  [Visitor economy 8](https://www.gov.uk/guidance/working-safely-during-coronavirus-covid-19/the-visitor-economy#shops-8) |
|  | Encouraging increased handwashing and introducing more handwashing facilities for workers handling goods and merchandise, or providing hand sanitiser where this is not practical. | How are you encouraging handwashing for workers handling goods or merchandise? | [Hotels and guest accommodation 5.4](https://www.gov.uk/guidance/working-safely-during-coronavirus-covid-19/hotels-and-other-guest-accommodation#section-5-4)  [Restaurants, pubs, bars and takeaway services 5.6](https://www.gov.uk/guidance/working-safely-during-coronavirus-covid-19/restaurants-offering-takeaway-or-delivery#takeaways-5-6)  [Food businesses](https://www.gov.uk/government/publications/covid-19-guidance-for-food-businesses/guidance-for-food-businesses-on-coronavirus-covid-19)  [Visitor economy 5.3](https://www.gov.uk/guidance/working-safely-during-coronavirus-covid-19/the-visitor-economy#shops-5-3)  [Visitor economy 8](https://www.gov.uk/guidance/working-safely-during-coronavirus-covid-19/the-visitor-economy#shops-8) |
| Inbound and outbound goods | Adjusting the way things are brought into the building and put away to create space for social distancing. | How have you altered the way things are brought into the building and stored to allow for social distancing? | [Hotels and guest accommodation 8](https://www.gov.uk/guidance/working-safely-during-coronavirus-covid-19/hotels-and-other-guest-accommodation#section-8)  [Restaurants, pubs, bars and takeaway services 8](https://www.gov.uk/guidance/working-safely-during-coronavirus-covid-19/restaurants-offering-takeaway-or-delivery#takeaways-8)  [Visitor economy 8](https://www.gov.uk/guidance/working-safely-during-coronavirus-covid-19/the-visitor-economy#shops-8) |
|  | Using non-contact deliveries where the nature of the product allows for use of electronic pre-booking. | Have you altered delivery procedures to create non-contact deliveries? | [Hotels and guest accommodation 8](https://www.gov.uk/guidance/working-safely-during-coronavirus-covid-19/hotels-and-other-guest-accommodation#section-8)  [Restaurants, pubs, bars and takeaway services 8](https://www.gov.uk/guidance/working-safely-during-coronavirus-covid-19/restaurants-offering-takeaway-or-delivery#takeaways-8)  [Visitor economy 5.4](https://www.gov.uk/guidance/working-safely-during-coronavirus-covid-19/the-visitor-economy#shops-5-4)  [Visitor economy 7.2](https://www.gov.uk/guidance/working-safely-during-coronavirus-covid-19/the-visitor-economy#shops-7-2)  [Visitor economy 8](https://www.gov.uk/guidance/working-safely-during-coronavirus-covid-19/the-visitor-economy#shops-8) |
|  | Creating one-way flow of traffic in stockrooms. | Have you created a one-way flow of traffic in stockrooms? | [Hotels and guest accommodation 8](https://www.gov.uk/guidance/working-safely-during-coronavirus-covid-19/hotels-and-other-guest-accommodation#section-8)  [Restaurants, pubs, bars and takeaway services 8](https://www.gov.uk/guidance/working-safely-during-coronavirus-covid-19/restaurants-offering-takeaway-or-delivery#takeaways-8)  [Visitor economy 5.4](https://www.gov.uk/guidance/working-safely-during-coronavirus-covid-19/the-visitor-economy#shops-5-4)  [Visitor economy 8](https://www.gov.uk/guidance/working-safely-during-coronavirus-covid-19/the-visitor-economy#shops-8) |
|  | Revising pick-up and drop-off collection points, procedures, signage and markings. | Have you provided signage for pick-up and drop-off collection points? | [Hotels and guest accommodation 8](https://www.gov.uk/guidance/working-safely-during-coronavirus-covid-19/hotels-and-other-guest-accommodation#section-8)  [Restaurants, pubs, bars and takeaway services 8](https://www.gov.uk/guidance/working-safely-during-coronavirus-covid-19/restaurants-offering-takeaway-or-delivery#takeaways-8)  [Visitor economy 5.4](https://www.gov.uk/guidance/working-safely-during-coronavirus-covid-19/the-visitor-economy#shops-5-4)  [Visitor economy 7.1](https://www.gov.uk/guidance/working-safely-during-coronavirus-covid-19/the-visitor-economy#shops-7-1)  [Visitor economy 8](https://www.gov.uk/guidance/working-safely-during-coronavirus-covid-19/the-visitor-economy#shops-8) |
|  | Minimising unnecessary contact at gatehouse security, yard and warehouse. | How have you minimised unnecessary contact at security, yard and warehouses? | [Hotels and guest accommodation 8](https://www.gov.uk/guidance/working-safely-during-coronavirus-covid-19/hotels-and-other-guest-accommodation#section-8)  [Restaurants, pubs, bars and takeaway services 8](https://www.gov.uk/guidance/working-safely-during-coronavirus-covid-19/restaurants-offering-takeaway-or-delivery#takeaways-8)  [Food businesses](https://www.gov.uk/government/publications/covid-19-guidance-for-food-businesses/guidance-for-food-businesses-on-coronavirus-covid-19)  [Visitor economy 8](https://www.gov.uk/guidance/working-safely-during-coronavirus-covid-19/the-visitor-economy#shops-8) |
|  | Considering methods to reduce frequency of deliveries, for example by ordering larger quantities less often. | How have you reduced frequency of deliveries? | [Hotels and guest accommodation 8](https://www.gov.uk/guidance/working-safely-during-coronavirus-covid-19/hotels-and-other-guest-accommodation#section-8)  [Restaurants, pubs, bars and takeaway services 8](https://www.gov.uk/guidance/working-safely-during-coronavirus-covid-19/restaurants-offering-takeaway-or-delivery#takeaways-8)  [Visitor economy 8](https://www.gov.uk/guidance/working-safely-during-coronavirus-covid-19/the-visitor-economy#shops-8) |
|  | Where possible and safe, having single workers load or unload vehicles. | Have you reduced loading or unloading to single workers where possible? | [Hotels and guest accommodation 8](https://www.gov.uk/guidance/working-safely-during-coronavirus-covid-19/hotels-and-other-guest-accommodation#section-8)  [Restaurants, pubs, bars and takeaway services 8](https://www.gov.uk/guidance/working-safely-during-coronavirus-covid-19/restaurants-offering-takeaway-or-delivery#takeaways-8)  [Visitor economy 8](https://www.gov.uk/guidance/working-safely-during-coronavirus-covid-19/the-visitor-economy#shops-8) |
|  | Maintaining consistent pairing where two-person deliveries are required. | Have you ensured consistent pairings where two-person deliveries are required? | [Visitor economy 7.2](https://www.gov.uk/guidance/working-safely-during-coronavirus-covid-19/the-visitor-economy#shops-7-2) |
|  | Encouraging drivers to stay in their vehicles where this does not compromise their safety and existing safe working practice. | Have you ensured that drivers stay in their vehicles, where this is safe? | [Hotels and guest accommodation 8](https://www.gov.uk/guidance/working-safely-during-coronavirus-covid-19/hotels-and-other-guest-accommodation#section-8)  [Restaurants, pubs, bars and takeaway services 8](https://www.gov.uk/guidance/working-safely-during-coronavirus-covid-19/restaurants-offering-takeaway-or-delivery#takeaways-8)  [Visitor economy 8](https://www.gov.uk/guidance/working-safely-during-coronavirus-covid-19/the-visitor-economy#shops-8) |

### 10 COVID risk domain: Healthcare

This section applies only where healthcare is provided as part of the event, e.g. in first-aid tents.

#### 10.1 General healthcare

| **Key considerations** | **Potential mitigations** | **Suggested questions** | **Relevant guidance** |
| --- | --- | --- | --- |
| Treatment of injuries | Where possible, all contact with members of the public should be carried out while maintaining social distancing measures. Where this is not possible, the principles for the Hierarchy of Risk should be applied*. | How have you ensured that healthcare will be provided whilst maintaining social distancing? | [Guidance for first responders](https://www.gov.uk/government/publications/novel-coronavirus-2019-ncov-interim-guidance-for-first-responders/interim-guidance-for-first-responders-and-others-in-close-contact-with-symptomatic-people-with-potential-2019-ncov) |
|  | Injuries occurring during the event should be treated. | - | [Return of sport](https://www.gov.uk/government/publications/coronavirus-covid-19-guidance-on-phased-return-of-sport-and-recreation/guidance-for-the-public-on-the-phased-return-of-outdoor-sport-and-recreation) |
| General protection of healthcare workers | After contact with an injured participant or member of the public, clean your hands thoroughly with soap and water or alcohol hand sanitiser at the earliest opportunity. Avoid touching your mouth, eyes and nose. | Please provide information on the guidance provided to healthcare workers around cleaning procedures following contact with the public. | [Return of sport](https://www.gov.uk/government/publications/coronavirus-covid-19-guidance-on-phased-return-of-sport-and-recreation/guidance-for-the-public-on-the-phased-return-of-outdoor-sport-and-recreation)  [Guidance for first responders](https://www.gov.uk/government/publications/novel-coronavirus-2019-ncov-interim-guidance-for-first-responders/interim-guidance-for-first-responders-and-others-in-close-contact-with-symptomatic-people-with-potential-2019-ncov) |
|  | There are no additional precautions to be taken in relation to cleaning your clothing or uniform other than what is usual practice. | - | [Return of sport](https://www.gov.uk/government/publications/coronavirus-covid-19-guidance-on-phased-return-of-sport-and-recreation/guidance-for-the-public-on-the-phased-return-of-outdoor-sport-and-recreation)  [Guidance for first responders](https://www.gov.uk/government/publications/novel-coronavirus-2019-ncov-interim-guidance-for-first-responders/interim-guidance-for-first-responders-and-others-in-close-contact-with-symptomatic-people-with-potential-2019-ncov) |
| Impact on local services | Undertake, in conjunction with local NHS services, detailed and continuous assessments to ensure there are no detrimental impacts of staging the event on the wider community and healthcare systems. | Have you undertaken assessments in conjunction with local NHS services to ensure no detrimental impacts of your event? | [Return of sport](https://www.gov.uk/government/publications/coronavirus-covid-19-guidance-on-phased-return-of-sport-and-recreation/guidance-for-the-public-on-the-phased-return-of-outdoor-sport-and-recreation) |

*Further details available in the [guidance for first responders](https://www.gov.uk/government/publications/novel-coronavirus-2019-ncov-interim-guidance-for-first-responders/interim-guidance-for-first-responders-and-others-in-close-contact-with-symptomatic-people-with-potential-2019-ncov).

#### 10.2 Cleaning following suspected or confirmed COVID cases in healthcare settings

| **Key considerations** | **Potential mitigations** | **Suggested questions** | **Relevant guidance** |
| --- | --- | --- | --- |
| Cleaning and disinfection | See “cleaning following suspected or confirmed COVID cases” section below. | As per relevant section. | As per relevant section. |

### 11 COVID risk domain: Performances and functions

This section applies only if the event involves the performing arts (e.g. concerts, music festivals, theatres). Please note that, in addition to the guidance below, the [Music Festivals supplementary guidance](https://www.thepurpleguide.co.uk/images/attachments/music-festivals-covid-19-supplementary-guidance-v1-13th-october-2020.pdf) from the Events Industry Forum also outlines similar mitigations.

#### 11.1 Location of performances and functions

| **Key considerations** | **Potential mitigations** | **Suggested questions** | **Relevant guidance** |
| --- | --- | --- | --- |
| Ensuring activity takes place outside wherever possible, including performance | Considering using available spaces outdoors for performances with a live audience in attendance. | If a live audience is in attendance, have you moved events outdoors where possible? | [Performing arts 3.2](https://www.gov.uk/guidance/working-safely-during-coronavirus-covid-19/performing-arts#arts-3-2) |
|  | Adapting live performing arts to ensure they are safe. If that is not possible, consider the use of technology solutions to reduce interactions and ensure social distancing. | How have you adapted live performing arts to ensure COVID security? | [Performing arts 4.1](https://www.gov.uk/guidance/working-safely-during-coronavirus-covid-19/performing-arts#arts-4-1) |
| Modification of indoor spaces | Taking steps to improve ventilation as far as possible, both through the use of mechanical systems and opening windows and doors. | How have you improved ventilation in indoor spaces? | [Performing arts 4.1](https://www.gov.uk/guidance/working-safely-during-coronavirus-covid-19/performing-arts#arts-4-1) |
|  | If working indoors, limiting the numbers to safely match the available ventilation of the space and the ability to observe social distancing. | How have you limited numbers to match the ventilation of the space? | [Performing arts 4.1](https://www.gov.uk/guidance/working-safely-during-coronavirus-covid-19/performing-arts#arts-4-1) |

#### 11.2 Performers

| **Key considerations** | **Potential mitigations** | **Suggested questions** | **Relevant guidance** |
| --- | --- | --- | --- |
| Limiting the number of performers as far as possible | Limiting the number of performers as far as possible. | Have you limited the number of performers? | [Performing arts introduction](https://www.gov.uk/guidance/working-safely-during-coronavirus-covid-19/performing-arts)  [Performing arts 4.1](https://www.gov.uk/guidance/working-safely-during-coronavirus-covid-19/performing-arts#arts-4-1) |
| Limiting the duration of performances as far as possible | Scheduling sufficient time between performances to reduce the possibility of different audiences coming into close proximity. | Have you scheduled time between performances to minimise mixing of audiences? | [Performing arts 3.11](https://www.gov.uk/guidance/working-safely-during-coronavirus-covid-19/performing-arts#arts-3-11)  [Performing arts 5.5](https://www.gov.uk/guidance/working-safely-during-coronavirus-covid-19/performing-arts#arts-5-5) |

#### 11.3 Performance attendees

| **Key considerations** | **Potential mitigations** | **Suggested questions** | **Relevant guidance** |
| --- | --- | --- | --- |
| Facilitating social distancing during performances | Reconfiguring entertainment spaces to enable audience to be seated rather than standing. | Have you reconfigured spaces to allow audiences to be seated rather than standing? | [Performing arts 3.2](https://www.gov.uk/guidance/working-safely-during-coronavirus-covid-19/performing-arts#arts-2-1)  [Places of worship 4](https://www.gov.uk/government/publications/covid-19-guidance-for-the-safe-use-of-places-of-worship-during-the-pandemic-from-4-july/covid-19-guidance-for-the-safe-use-of-places-of-worship-during-the-pandemic-from-4-july)  [Visitor economy 2.2](https://www.gov.uk/guidance/working-safely-during-coronavirus-covid-19/the-visitor-economy#shops-4-2) |
| Interaction between attendees | Providing clear communication, demarcating spaces, using sufficient ushers. | How have you demarcated spaces to allow for social distancing? | [Performing arts 3.2](https://www.gov.uk/guidance/working-safely-during-coronavirus-covid-19/performing-arts#arts-2-1)  [Visitor economy 2.2](https://www.gov.uk/guidance/working-safely-during-coronavirus-covid-19/the-visitor-economy#shops-4-2) |
|  | Avoiding gatherings that may encourage audience behaviours that increase physical contact outside of household groups or support bubbles. | How have you minimised the risk of audience gatherings outside of household groups or support bubbles? | [Performing arts 3.2](https://www.gov.uk/guidance/working-safely-during-coronavirus-covid-19/performing-arts#arts-2-1)  [Performing arts 3.4](https://www.gov.uk/guidance/working-safely-during-coronavirus-covid-19/performing-arts#arts-3-4)  [Places of worship 4](https://www.gov.uk/government/publications/covid-19-guidance-for-the-safe-use-of-places-of-worship-during-the-pandemic-from-4-july/covid-19-guidance-for-the-safe-use-of-places-of-worship-during-the-pandemic-from-4-july) |
| Seating | Audiences should be seated as individuals or groups from the same household or support bubble. | Have you ensured audiences are seated as individuals or as households/support bubbles? | [Performing arts 3.12](https://www.gov.uk/guidance/working-safely-during-coronavirus-covid-19/performing-arts#arts-3-12)  [Places of worship 4](https://www.gov.uk/government/publications/covid-19-guidance-for-the-safe-use-of-places-of-worship-during-the-pandemic-from-4-july/covid-19-guidance-for-the-safe-use-of-places-of-worship-during-the-pandemic-from-4-july) |
|  | Providing allocated seating and managing seating plans through ticketing systems. | Have you provided allocated seating? | [Performing arts 3.12](https://www.gov.uk/guidance/working-safely-during-coronavirus-covid-19/performing-arts#arts-3-12) |
|  | Installing seat separation or labelling seats which should not be used. | How have you labelled seats which should not be used? | [Performing arts 3.12](https://www.gov.uk/guidance/working-safely-during-coronavirus-covid-19/performing-arts#arts-3-12) |
|  | Deploying staff to support the audience in adhering to social distanced seating. | Have you deployed staff to support the audience in social distancing? | [Performing arts 3.12](https://www.gov.uk/guidance/working-safely-during-coronavirus-covid-19/performing-arts#arts-3-12) |
|  | Venues with balconies should keep the front 2m of seats empty to minimise aerosol risk to seated attendees. | If you have balconies, have the first 2m of these been kept empty? | [Performing arts 3.2](https://www.gov.uk/guidance/working-safely-during-coronavirus-covid-19/performing-arts#arts-2-1) |
|  | Considering the needs of disabled audience members, for example access to captioning or audio description services, when managing seating. | How have you considered the needs of disabled audience members when managing seating? | [Performing arts 3.12](https://www.gov.uk/guidance/working-safely-during-coronavirus-covid-19/performing-arts#arts-3-12) |

#### 11.4 Performance and function spaces

| **Key considerations** | **Potential mitigations** | **Suggested questions** | **Relevant guidance** |
| --- | --- | --- | --- |
| Minimising cross-contamination between performance attendees | Cleaning auditoria very frequently, typically between each performance. Scheduling performances to allow sufficient time to undertake necessary cleaning before the next audience arrives with particular attention paid to surfaces that hands of audience and staff are likely to come into contact with such as doors, seat arms and handrails. | Please detail procedures and frequency for cleaning of auditoria and function spaces. | [Performing arts 3.12](https://www.gov.uk/guidance/working-safely-during-coronavirus-covid-19/performing-arts#arts-3-12)  [Performing arts 5.5](https://www.gov.uk/guidance/working-safely-during-coronavirus-covid-19/performing-arts#arts-5-5) |

### 12 COVID risk domain: Sporting events

This section applies only to sporting events (both spectator sporting events and those with large-group participation e.g. marathons).

#### 12.1 Spectators

| **Key considerations** | **Potential mitigations** | **Suggested questions** | **Relevant guidance** |
| --- | --- | --- | --- |
| Risk assessment | Where it is anticipated that an activity will attract spectators, there should be a named person or persons with responsibility for ensuring adherence with these guidelines and ensuring the facility is COVID19 Secure. | Have you named a person responsible for ensuring that spectator sporting events are COVID-secure? | [Return of sport](https://www.gov.uk/government/publications/coronavirus-covid-19-guidance-on-phased-return-of-sport-and-recreation/guidance-for-the-public-on-the-phased-return-of-outdoor-sport-and-recreation) |

#### 12.2 Participants

| **Key considerations** | **Potential mitigations** | **Suggested questions** | **Relevant guidance** |
| --- | --- | --- | --- |
| Risk awareness | Make participants aware of any increased risk associated with taking part in activity, based on the assessment undertaken by the event organiser. | Have you made participants aware of increased risk associated with participating in the event? | [Return of sport](https://www.gov.uk/government/publications/coronavirus-covid-19-guidance-on-phased-return-of-sport-and-recreation/guidance-for-the-public-on-the-phased-return-of-outdoor-sport-and-recreation) |
|  | Advise participants to comply with public health restrictions and avoid high risk behaviour outside the sports setting | Have you advised participants to comply with public health restrictions? | [Return of sport](https://www.gov.uk/government/publications/coronavirus-covid-19-guidance-on-phased-return-of-sport-and-recreation/guidance-for-the-public-on-the-phased-return-of-outdoor-sport-and-recreation) |
| Minimising transmission risk | Participants should be discouraged from bringing any equipment, baggage, or clothing that is not essential for their participation in the event. | Have you advised participants not to bring any non-essential equipment, baggage or clothing to the event? | [Return of sport](https://www.gov.uk/government/publications/coronavirus-covid-19-guidance-on-phased-return-of-sport-and-recreation/guidance-for-the-public-on-the-phased-return-of-outdoor-sport-and-recreation) |

#### 12.3 The course

| **Key considerations** | **Potential mitigations** | **Suggested questions** | **Relevant guidance** |
| --- | --- | --- | --- |
| Facilitating social distancing during events | Ensure that pre-start assembly areas, the start line and holding areas are designed so that participants do not need to assemble at the start of the event in a manner which conflicts with social distancing guidelines. | How have you ensured social distancing at pre-start assembly, assembly and holding areas? | [Return of sport](https://www.gov.uk/government/publications/coronavirus-covid-19-guidance-on-phased-return-of-sport-and-recreation/guidance-for-the-public-on-the-phased-return-of-outdoor-sport-and-recreation) |
|  | Consider rolling start times to allow social distancing to be maintained. | Have you considered rolling start times, if possible? | [Return of sport](https://www.gov.uk/government/publications/coronavirus-covid-19-guidance-on-phased-return-of-sport-and-recreation/guidance-for-the-public-on-the-phased-return-of-outdoor-sport-and-recreation) |
|  | Capacity and density of the participants on the course should always allow for social distancing. | How have you ensured social distancing of participants on the course? | [Return of sport](https://www.gov.uk/government/publications/coronavirus-covid-19-guidance-on-phased-return-of-sport-and-recreation/guidance-for-the-public-on-the-phased-return-of-outdoor-sport-and-recreation) |
|  | Consider pinch points on the course before, during and after the event and manage them accordingly | How have you managed pinch points on the course, if appropriate? | [Return of sport](https://www.gov.uk/government/publications/coronavirus-covid-19-guidance-on-phased-return-of-sport-and-recreation/guidance-for-the-public-on-the-phased-return-of-outdoor-sport-and-recreation) |
|  | Features such as entertainment should be withdrawn. | Have you withdrawn features such as entertainment? | [Return of sport](https://www.gov.uk/government/publications/coronavirus-covid-19-guidance-on-phased-return-of-sport-and-recreation/guidance-for-the-public-on-the-phased-return-of-outdoor-sport-and-recreation) |
|  | Potential contact points such as the handling of medals, timing chips and numbers should be managed appropriately. | How have you managed contact points e.g. handling of medals or timing chips? | [Return of sport](https://www.gov.uk/government/publications/coronavirus-covid-19-guidance-on-phased-return-of-sport-and-recreation/guidance-for-the-public-on-the-phased-return-of-outdoor-sport-and-recreation) |
|  | Access to feed and drink stations should be provided in such a way that social distancing can still be observed by officials and participants. | How have you ensured that feed and drink stations ensure social distancing, if these are provided? | [Return of sport](https://www.gov.uk/government/publications/coronavirus-covid-19-guidance-on-phased-return-of-sport-and-recreation/guidance-for-the-public-on-the-phased-return-of-outdoor-sport-and-recreation) |

### 13 COVID risk domain: Ceremonies and religious events

Please note that specific capacity limitations apply for religious events, as outlined in the [places of worship guidance](https://www.gov.uk/government/publications/covid-19-guidance-for-the-safe-use-of-places-of-worship-during-the-pandemic-from-4-july/covid-19-guidance-for-the-safe-use-of-places-of-worship-during-the-pandemic-from-4-july) section 5. For all others, capacity considerations apply as outlined in COVID risk domain 1.

Religious events with singing or chanting should consult the performing arts guidance. Post-religious functions (e.g. wedding receptions) should comply with the other sections of the framework.

This section does not apply to childcare taking place in a place of worship. For this, please consult the above guidance.

#### 13.1 Event-specific capacity limits

| **Key considerations** | **Potential mitigations** | **Suggested questions** | **Relevant guidance** |
| --- | --- | --- | --- |
| Capacity limits | Marriage ceremonies, commemorative ceremonies, support groups should have no more than 15 people; funerals no more than 30 people | If your event is a marriage ceremony, commemorative ceremony, support group or funeral, have you adhered to the specific capacity limit? | [Marriages](https://www.gov.uk/government/publications/covid-19-guidance-for-small-marriages-and-civil-partnerships/covid-19-guidance-for-small-marriages-and-civil-partnerships)  [Places of worship](https://www.gov.uk/government/publications/covid-19-guidance-for-the-safe-use-of-places-of-worship-during-the-pandemic-from-4-july/covid-19-guidance-for-the-safe-use-of-places-of-worship-during-the-pandemic-from-4-july) |

#### 13.2 Adaptations to services

| **Key considerations** | **Potential mitigations** | **Suggested questions** | **Relevant guidance** |
| --- | --- | --- | --- |
| Service length | Ceremonies and services should be concluded in the shortest reasonable time. | Is your ceremony or service taking place in the shortest period of time possible? | [Marriages](https://www.gov.uk/government/publications/covid-19-guidance-for-small-marriages-and-civil-partnerships/covid-19-guidance-for-small-marriages-and-civil-partnerships)  [Places of worship 4](https://www.gov.uk/government/publications/covid-19-guidance-for-the-safe-use-of-places-of-worship-during-the-pandemic-from-4-july/covid-19-guidance-for-the-safe-use-of-places-of-worship-during-the-pandemic-from-4-july) |
| Changes to service | Stream worship or other events to avoid large gatherings and to continue to reach those individuals who are self-isolating or particularly vulnerable to COVID-19. | Have you considered live-streaming events to minimise risk of large gatherings? | [Marriages](https://www.gov.uk/government/publications/covid-19-guidance-for-small-marriages-and-civil-partnerships/covid-19-guidance-for-small-marriages-and-civil-partnerships)  [Places of worship 4](https://www.gov.uk/government/publications/covid-19-guidance-for-the-safe-use-of-places-of-worship-during-the-pandemic-from-4-july/covid-19-guidance-for-the-safe-use-of-places-of-worship-during-the-pandemic-from-4-july) |
|  | Where food or drink (‘consumables’) are essential to the act of worship, they can be used, however the sharing of food should be avoided, as should the use of communal vessels. | Have you removed any food sharing or communal vessels from the ceremony? | [Marriages](https://www.gov.uk/government/publications/covid-19-guidance-for-small-marriages-and-civil-partnerships/covid-19-guidance-for-small-marriages-and-civil-partnerships)  [Places of worship 4](https://www.gov.uk/government/publications/covid-19-guidance-for-the-safe-use-of-places-of-worship-during-the-pandemic-from-4-july/covid-19-guidance-for-the-safe-use-of-places-of-worship-during-the-pandemic-from-4-july) |
|  | Consumables should be securely covered, and prior to the receptacle being opened, it should be cleaned, hands should be washed or gloves worn. | Have you ensured any consumables are securely covered prior to consumption? | [Marriages](https://www.gov.uk/government/publications/covid-19-guidance-for-small-marriages-and-civil-partnerships/covid-19-guidance-for-small-marriages-and-civil-partnerships)  [Places of worship 4](https://www.gov.uk/government/publications/covid-19-guidance-for-the-safe-use-of-places-of-worship-during-the-pandemic-from-4-july/covid-19-guidance-for-the-safe-use-of-places-of-worship-during-the-pandemic-from-4-july) |
|  | Any pre-requisite washing/ablution rituals should not be done at the place of worship but carried out prior to arrival. | Have you removed any washing or ablution rituals? | [Marriages](https://www.gov.uk/government/publications/covid-19-guidance-for-small-marriages-and-civil-partnerships/covid-19-guidance-for-small-marriages-and-civil-partnerships)  [Places of worship 4](https://www.gov.uk/government/publications/covid-19-guidance-for-the-safe-use-of-places-of-worship-during-the-pandemic-from-4-july/covid-19-guidance-for-the-safe-use-of-places-of-worship-during-the-pandemic-from-4-july) |
| Cash donations | Where possible faith leaders should discourage cash donations and continue to use online or contactless giving and resources.  Where this is not an option, cash should be collected in a receptacle that is set in one place and handled by one individual, as opposed to being passed around. | Have you made changes to cash donation systems to be contactless where possible? | [Places of worship 4](https://www.gov.uk/government/publications/covid-19-guidance-for-the-safe-use-of-places-of-worship-during-the-pandemic-from-4-july/covid-19-guidance-for-the-safe-use-of-places-of-worship-during-the-pandemic-from-4-july) |

### 14 Local considerations

Although not linked to specific articles of national guidance, other considerations should be made to the context of the region in which the event is taking place. These have been agreed based on expert views and consensus. They fall into two broad categories: considerations relating to the event specifically, but not mentioned in the national guidance, and considerations of the local and national context in which these events are taking place.

#### 14.1 Event-related considerations

These are considerations which have been recommended by colleagues responsible to either licensing the event and or ensuring that it is COVID secure as being valuable in ensuring COVID-security but do not have a direct link to national guidance.

These considerations include:

- Asking event organisers to provide the region of origin of attendees and staff (including contractors) to assess risk of widespread transmission or cross-region spread
- Ensuring that no one attends from areas in tiers 2 or 3 in the three-tier government local COVID alert level
- Assessing the risks posed to protected groups, or the risk of regional or national spread
- Ensuring that events have a clear policy outlining refund procedures in case of COVID-related issues (to facilitate attendee adherence to public health guidance e.g. staying home if they have symptoms consistent with COVID-19)

Please note that contractors are the responsibility of the event organiser; it is the responsibility of the organiser to provide evidence that the contractors are adhering to COVID-secure guidelines. It may be beneficial to include this requirement in the contracts between organisers and external organisations.

#### 14.2 Local and national context

Considerations which relate to the local and national context are intended to account for the fact that these events do not take place in isolation, and that although an event may itself be COVID-secure, there may be external considerations which may impact on this. An extreme example would be if a local lockdown were imposed, prohibiting large gatherings.

These considerations include:

- Gathering disease prevalence and transmission rates in the area
- Assessing the cumulative risk of multiple large events taking place within a short space of time
- Current and predicted trend in number of COVID cases and impact on the local and national NHS

These are summarised, in addition to restrictions placed on areas with heightened COVID alert levels, in the [Local COVID alert levels guidance](https://www.gov.uk/guidance/local-covid-alert-levels-what-you-need-to-know).

These considerations are in addition to the national guidance highlighted in the previous sections. The outcome of the checklist (derived from the framework) should be used in conjunction with the information gathered at a local level to inform recommendations on events.

### 15 Appendix 1: List of included guidance

**Guidance from GOV.UK**

- [COVID-19: cleaning in non-healthcare settings outside the home](https://www.gov.uk/government/publications/covid-19-decontamination-in-non-healthcare-settings/covid-19-decontamination-in-non-healthcare-settings)
- [COVID-19: guidance for first responders](https://www.gov.uk/government/publications/novel-coronavirus-2019-ncov-interim-guidance-for-first-responders/interim-guidance-for-first-responders-and-others-in-close-contact-with-symptomatic-people-with-potential-2019-ncov)
- [COVID-19: Guidance for small marriages and civil partnerships](https://www.gov.uk/government/publications/covid-19-guidance-for-small-marriages-and-civil-partnerships/covid-19-guidance-for-small-marriages-and-civil-partnerships)
- [COVID-19: Guidance for the safe use of multi-purpose community facilities](https://www.gov.uk/government/publications/covid-19-guidance-for-the-safe-use-of-multi-purpose-community-facilities/covid-19-guidance-for-the-safe-use-of-multi-purpose-community-facilities)
- [COVID-19: guidance for the safe use of places of worship and special religious services and gatherings during the pandemic](https://www.gov.uk/government/publications/covid-19-guidance-for-the-safe-use-of-places-of-worship-during-the-pandemic-from-4-july)
- [COVID-19: guidance for the safe use of places of worship during the pandemic](https://www.gov.uk/government/publications/covid-19-guidance-for-the-safe-use-of-places-of-worship-during-the-pandemic-from-4-july/covid-19-guidance-for-the-safe-use-of-places-of-worship-during-the-pandemic-from-4-july)
- [Guidance for food businesses on coronavirus (COVID-19)](https://www.gov.uk/government/publications/covid-19-guidance-for-food-businesses/guidance-for-food-businesses-on-coronavirus-covid-19)
- [Guidance for the public on the phased return of outdoor sport and recreation in England](https://www.gov.uk/government/publications/coronavirus-covid-19-guidance-on-phased-return-of-sport-and-recreation/guidance-for-the-public-on-the-phased-return-of-outdoor-sport-and-recreation)
- [COVID-19: Guidance for wedding and civil partnership receptions and celebrations](https://www.gov.uk/government/publications/covid-19-guidance-for-small-marriages-and-civil-partnerships/covid-19-guidance-for-wedding-and-civil-partnership-receptions-and-celebrations)
- [Local COVID alert levels guidance](https://www.gov.uk/guidance/local-covid-alert-levels-what-you-need-to-know)
- [Maintaining records of staff, customers and visitors to support NHS Test and Trace](https://www.gov.uk/guidance/maintaining-records-of-staff-customers-and-visitors-to-support-nhs-test-and-trace)
- [Working safely during coronavirus (COVID-19): Hotels and other guest accommodation](https://www.gov.uk/guidance/working-safely-during-coronavirus-covid-19/hotels-and-other-guest-accommodation)
- [Working safely during coronavirus (COVID-19): Performing arts](https://www.gov.uk/guidance/working-safely-during-coronavirus-covid-19/performing-arts)
- [Working safely during coronavirus (COVID-19): Restaurants, pubs, bars and takeaway services](https://www.gov.uk/guidance/working-safely-during-coronavirus-covid-19/restaurants-offering-takeaway-or-delivery)
- [Working safely during coronavirus (COVID-19): The visitor economy](https://www.gov.uk/guidance/working-safely-during-coronavirus-covid-19/the-visitor-economy)

**Guidance from other bodies**

- [Keeping workers and audiences safe during COVID-19 In the Outdoor Event Industry in England](https://www.eventindustrynews.com/wp-content/uploads/2020/07/EIfDCMS-COVID-19-Working-Safely-9-July-2020.pdf) (Events Industry Forum)
- [Music Festivals – COVID-19 Supplementary Guidance](https://www.thepurpleguide.co.uk/images/attachments/music-festivals-covid-19-supplementary-guidance-v1-13th-october-2020.pdf) (Events Industry Forum)
- [COVID-19: outdoor events guidance](https://www.local.gov.uk/covid-19-outdoor-events-guidance) (Local Government Association)
- [Guide to Safety at Sports Grounds Supplementary Guidance 02: Planning for social distancing at sports grounds](https://sgsa.org.uk/wp-content/uploads/2020/08/SG02-Planning-for-Social-Distancing-at-Sports-Grounds.pdf) (Sports Grounds Safety Authority)
- [Don’t rely on temperature screening products for detection of coronavirus (COVID-19), says MHRA](https://www.gov.uk/government/news/dont-rely-on-temperature-screening-products-for-detection-of-coronavirus-covid-19-says-mhra)
