## Appendix 3: C-SET user manual for "The COVID-19 Suffolk Events Toolkit (C-SET): A structured approach to conducting COVID-secure events"

**C-SET CHECKLIST – USER MANUAL**

***Introduction***

- This checklist is based on the “C-SET” PHS framework. Compared to the framework, the checklist has a condensed amount of questions for ease of use, and some questions may refer to several areas of the framework. The checklist is not exhaustive and of general nature – for full details refer to the framework.
- The checklist is aimed at helping event organisers, Safety Advisory Groups, and Public Health teams determine if an event has the necessary COVID19-specific risk mitigations in place.

***Notes on completion – providing the evidence from event organiser***

- Work your way through the checklist, starting with the event profile and then all the relevant checklist sections (as you fill the event profile and answer the screening questions, it will be indicated which sections need completing).
- The supporting evidence column is free text - please keep this succinct. If you want to attach or embed any documents for more details, please clearly refer to the relevant page and section within the attached document that answers the specific question. For example, "as per attached document 1 page 2, section 2.3" and not just "see attached document".
- Please note that contractors are the responsibility of the event organiser; therefore, it is not sufficient to say “this is the contractor’s responsibility” in answer to any of the checklist questions. It is the responsibility of the organiser to provide evidence that the contractors are adhering to COVID-secure guidelines. It may be beneficial to include this requirement in the contracts between organisers and external organisations.
- The checklist is aimed to be comprehensive so some questions may not be applicable to your event. If so, please write “not applicable” against the question.

***Notes on completion – reviewing the evidence***

- For bodies assessing COVID-security using this framework, review the evidence gathered in the checklist. As you review the evidence, against each checklist question please fill:
  - RAG assessment – no significant concern/ minor concerns / major concerns (this is a built-in dropdown)
  - Brief rationale for the assessment
  - Any suggestions/recommendations for improvement
- Summary tab - a summary of areas of concern is automatically generated in the summary tab of the checklist. Domain- and subdomain- specific recommendations can then be made by referring to the full framework, to enable organisers to make positive changes to their events.
- Please note that this is not a quantitative tool to determine whether the event can go ahead or not. This tool helps to capture information in a comprehensive way so that appropriate improvements can be suggested and made. It is up to local systems to determine the best applications of this tool.
